# Public toilets have reduced enteric pathogen hazards in San Francisco

**DOI:** 10.1101/2023.02.10.23285757

**Authors:** Troy Barker, Drew Capone, Heather K. Amato, Ryan Clark, Abigail Henderson, David A. Holcomb, Elizabeth Kim, Jillian Pape, Emily Parker, Thomas VanderYacht, Jay Graham, Joe Brown

## Abstract

Uncontained fecal wastes in cities may present exposure risks to the public. We collected discarded feces from public spaces in San Francisco for analysis by RT-qPCR for a range of enteric pathogens. Out of 59 samples, we found 12 (20%) were of human origin and 47 (80%) were non-human; 30 of 59 stools were positive for ≥1 of the 35 pathogens assessed, including pathogenic *E. coli, Shigella*, norovirus, *Cryptosporidium*, and *Trichuris*. Using quantitative enteric pathogen estimates and data on observed fecal waste from a public reporting system, we modeled pathogens removed from the environment attributable to a recently implemented program of public toilet construction. We estimated that each new public toilet reduced the annual number of enteric pathogens released into the immediate environment (within 500 m walking distance), including 6.3 × 10^12^ enteropathogenic *E. coli* (95% CI: 4.0 × 10^12^ – 7.9 × 10^12^), 3.2 × 10^11^ enteroaggregative *E. coli* (95% CI: 1.3 × 10^11^ – 6.3 × 10^11^), and 3.2 × 10^8^ *Shigella* (6.3 × 10^7^ – 2.5 × 10^9^). Improving access to public sanitation can reduce enteric pathogen hazards in cities. Interventions must also consider the hygienic disposal of animal waste to reduce microbial hazards with zoonotic infection potential.

**SYNOPSIS:** This paper describes enteric pathogen hazards from discarded feces on the streets of San Francisco and estimates their reduction following a public toilet intervention.

**TOC/Abstract art:** Created with BioRender and a photograph by author Jay Graham

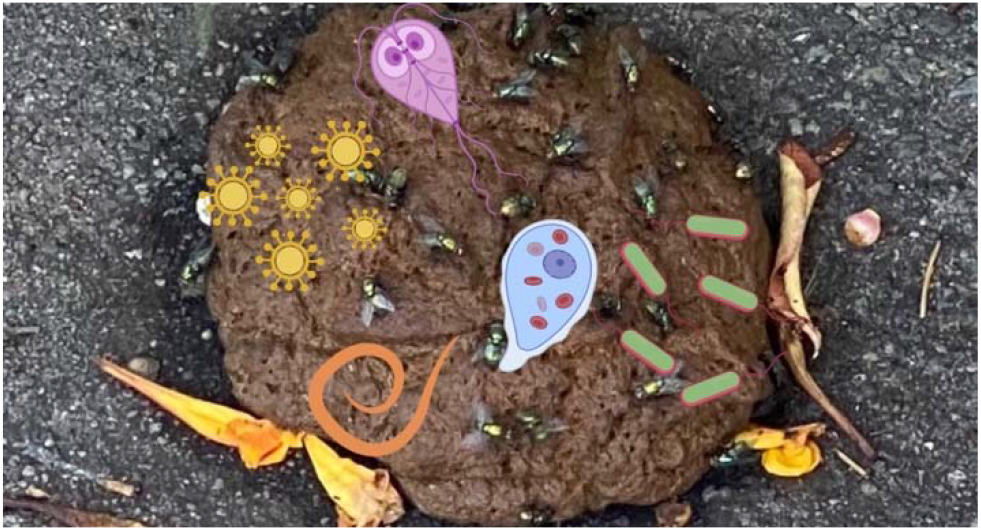

## INTRODUCTION

People experiencing homelessness are more likely than the general population to suffer from communicable diseases^1^, partly because they lack consistent access to basic infrastructure including safe water and sanitation^2, 3^. Nearly one million persons in US cities have insufficient access to basic sanitation and over 600,000 cannot consistently access basic water infrastructure^2^, primarily those lacking stable housing.

Nearly 130,000 people in California experience homelessness each day, and they are disproportionately unsheltered compared to those in other states^1, 4^. Like many US cities, San Francisco does not have enough public toilets to meet demand^5^. Consistent access to sanitation and soap and water for proper handwashing is necessary to prevent the spread of enteric pathogens that may cause diarrheal disease^6^. Without safe, hygienic, and publicly accessible toilets when and where people need them, open defecation is common^7-9^. As a result, uncontained fecal waste can accumulate near where people live, work, and play, creating opportunities for exposure to enteric pathogens through well-known direct and indirect pathways^10-13^.

Fecal contamination on the streets of San Francisco is so common that the Department of Public Works (DPW) created a well-used system to report feces in public spaces, where instances of fecal waste can be reported by dialing 311 from a telephone (or, now, via a website or Twitter)^11,14^. Data since 2008 are publicly available at https://datasf.org/opendata/. As a step toward addressing the well-publicized problem, DPW implemented the Pit Stop program beginning in 2014, aiming to reduce open defecation in public spaces by installing staffed public toilets in areas of high need.^11,12^ The intervention also includes animal waste bags and waste bins for disposal. A full description of the program and its history is available at the Pit Stop website: https://sfpublicworks.wixsite.com/pitstop. A recent impact assessment of the program estimated that the installation of public latrines reduced 311 reports of fecal waste on the street by a mean of 12.5 stools per week within 500 meters (walking distance) of newly installed Pit Stop locations in the six months following installation, compared with a pre-intervention baseline^15^. The greatest reduction in reports of fecal waste occurred in the Tenderloin neighborhood (i.e., 18 stools per week per new facility), which had 10 public toilets, the most of any neighborhood in the program. The Tenderloin and South of Market (SoMa) had the highest counts of 311 reports in the study period; together, these neighborhoods make up District 6, which had the highest count of people experiencing homelessness (n=3,656) in SF as of 2019^16^. With only 3 Pit Stop public facilities in SoMa, there was no significant reduction of 311 reports associated with the Pit Stop intervention in this neighborhood. A recent independent analysis by the San Francisco Chronicle of 311 data concluded that, while feces-related service requests for every other neighborhood in San Francisco have gone up an average of 400% in the period from 2012-2021, reports for the Tenderloin reduced by 29%, with pronounced further decreases in the area immediately around three Pit Stop interventions^14^.

These interventions may affect exposures to enteric pathogen hazards to the public from human and animal fecal waste, including among people experiencing homelessness who may bear the greatest direct risks. Our primary aim was to estimate the degree to which Pit Stop interventions have reduced enteric pathogens in the immediate environment surrounding new toilet facilities. To this end, we: (1) conducted a systematic survey of discarded feces in a pre-defined area; (2) determined whether each fecal sample was of human or non-human origin; (3) quantified a range of enteric pathogens in recovered fecal samples, using molecular methods; and (4) used the intervention-attributable reduction in observed feces from 311 reports^15^ to estimate reductions of the enteric pathogens we detected.

## MATERIALS AND METHODS

A previous longitudinal study estimated that the installation of public toilets in two San Francisco neighborhoods resulted in a mean reduction of 18 stools per week within 500 meters (walking distance) of each facility^15^. We systematically collected discarded stools in this area and quantified a range of enteric pathogens in each sample. We used these data to model the reduction of pathogen hazards attributable to each facility.

### Sample Collection

We used 311 reports of fecal waste in San Francisco, CA in August 2020 to identify hotspots for open defecation (OD) reports and design a systematic survey of discarded stools for enteric pathogen analysis. The available 311 data include reports of any discarded feces, including both suspected human and non-human fecal waste. The Tenderloin and SoMa neighborhoods had the highest number of 311 reports and were therefore selected for sampling; we matched observed feces with 311 reports to verify that 311 data reliably indicated instances of fecal waste (Text S1). We prioritized 20 blocks, including both sidewalks on either side of the street for collection (Figure S1, Figure S2). We generated a perimeter around the selected blocks, with any block inside the perimeter potentially utilized if samples were not available on the 20 selected blocks. We collected biospecimens on four Wednesday mornings in September and October of 2020 before street cleaning began. Stool samples were collected into one-liter biohazard bags and stored in a cooler with ice packs. We transferred samples into 1.5 mL cryotubes and stored them at -20°C within 4 hours of collection. Any confirmed animal stool (e.g., if the team observed an animal defecating) was not collected.

### Sample preparation and analysis

We used the QIAamp 96 Virus QIAcube HT Kit (Qiagen, Hilden, Germany) to extract nucleic acids from 100 mg of stool using a pre-treatment step previously validated for molecular detection of multiple enteric pathogens with both DNA and RNA genomes (Text S2)^17, 18^. We proceeded with extraction following the manufacturer’s protocol for the QIAamp 96 Virus QIAcube HT Kit, which we automated on the QIAcube (Qiagen, Hilden, Germany). We measured the concentration of dsDNA using the dsDNA HS assay with a Qubit 4 Fluorometer (Invitrogen, Waltham, Massachusetts).

We quantified human mitochondrial DNA (mtDNA) in each sample by dPCR to determine whether feces were of presumptive human origin (Text S5, Figure S3) using an assay that previously demonstrated 100% sensitivity and 97% specificity to human stool^19^. We normalized mtDNA gene copy estimates to ng of dsDNA, and compared results against values reported in the literature to categorize samples as human or non-human.^19^ Positive and negative PCR controls^20^ were run each day of analysis via qPCR (Text S3) and dPCR (Text S5, Table S7). We developed and used custom TaqMan Array Cards (TAC) (Thermo Fisher Scientific, Waltham, MA) using published primer and probe sequences (Tables S2-S4) for a range of enteric bacteria, viruses, protozoa, helminths, and controls. TAC is a 384-well array card with 8 ports for loading samples and each 1.5 μL well contains lyophilized primers and probes for the detection of pre-defined targets. We analyzed extracted nucleic acids via TAC on the QuantStudio 7, generating real-time RT-qPCR curves for each target for each sample (Text S3). Standard curve details and 95% limits of detection are presented in Table S3. We visually compared exponential curves and multicomponent plots with positive control plots to validate positive amplification^21, 22^. We manually set thresholds to the point of inflection and considered targets amplifying at or below 35 cycles positive^23^. We re-ran samples that did not amplify DNA/RNA extraction positive controls as expected at a 1:10 dilution, and samples that did not then amplify controls were excluded from analysis. We performed additional confirmatory analysis of samples positive for soil-transmitted helminths via microscopy using the mini-FLOTAC method (Text S4, Figure S4).^16^ We transformed cycle quantification (Cq) values into gene copy concentrations per gram feces using pathogen-specific standard curves.

### Stochastic model

We estimated the annual number of pathogens diverted from the environment attributable to the Pit Stop intervention program using R version 4.1.0^24^. We transformed gene copies (gc) into genomic units (i.e., discrete pathogens) using published values for gene copies per genome (Table S5, model parameters)^23^. We treated non-detects as true zeroes. Using the fitdistrplus package in R^25^, we used maximum likelihood estimation (MLE) to fit log-normal distributions to the quantity of pathogens per gram of stool in fecal samples, obtaining separate estimates of the mean and standard deviation of log-pathogens per gram of presumptively human and non-human stool, respectively, for each pathogen detected. We thus generated an estimated mean and standard deviation for the number of pathogens per gram of feces. However, we assigned upper bound thresholds to the distributions of pathogen concentrations in feces based on biological plausibility (i.e., bacteria: 10^9^/gram; viruses: 10^12^/gram; protozoa: 10^7^/gram; helminths: 10^6^/gram) and to prevent outliers in the distribution from driving the overall annual estimate. We modeled an estimate of 18 stools per week that were diverted from the environment^15^ due to each new toilet facility across 52 weeks, because this was the reduction reported by Amato *et al*. 2022^15^ for the Tenderloin and SOMA neighborhoods. Using a Monte Carlo Simulation, we applied a binomial distribution to estimate whether each unique diverted stool (of 18) contained a specific pathogen. For each stool simulated to contain a given pathogen, we estimated the concentration using the corresponding MLE-generated distribution. We fit a log-normal distribution to the mass of human defecation events reported in Cummings *et al*. 1992 to stochastically estimate the mass of each stool^26^. For pathogens detected in only one of the collected samples, we used the number of pathogens per gram of stool for the single stool in place of an MLE-generated mean and the average MLE standard deviation from plurally detected pathogens (Table S5). We repeated the process 52 times to estimate pathogens reduced over a year. We then summed the number of estimated pathogens across the entire year to estimate the number of pathogens diverted annually, repeating the process 1000 times to generate 95% confidence intervals.

## RESULTS

We tested 60 stool samples, but excluded one whose assays lacked positive control amplification, leaving 59 samples for analysis. Positive controls exhibited consistent amplification (Cq∼20) and no amplification was observed in our negative controls, except for the 16S assay (Cq∼35, Text S6), which is a known contaminant in mastermix containing the TaqMan polymerase^27,28^. Pathogen prevalence disaggregated by human mtDNA results is shown in Table 1; we determined that 12 samples were of human origin and 47 were from non-human sources. Out of 35 pathogens analyzed using TAC, 30/59 samples (51%) contained one or more pathogens, 21/59 (36%) contained two or more pathogens, 8/59 (14%) contained 3 or more pathogens, and 1/59 (2%) contained 5 pathogens. The most prevalent pathogens were atypical enteropathogenic *E. coli* (EPEC) at 37% (22/59), typical EPEC and *Acanthamoeba spp*. each at 12% (7/59), *Cryptosporidium* at 8% (5/59), and *Giardia* at 7% (4/59).

**Table 1.** Prevalence of pathogens in collected stool samples.

| Pathogen | All fecal samples<br>(n=59) | Presumptively<br>human<sup>e</sup> (n=12) | Presumptively non-human<sup>e</sup> (n=47) |
| --- | --- | --- | --- |
| **Bacteria** |  |  |  |
| EPEC<sup>a</sup> (atypical) | 37% | 42% | 36% |
| EPEC<sup>a</sup> (typical) | 12% | 0% | 15% |
| EAEC<sup>b</sup> (aaiC/aatA) | 3% | 8% | 2% |
| ETEC<sup>c</sup> (LT/ST) | 3% | 8% | 2% |
| *Helicobacter pylori* | 3% | 17% | 0% |
| *Shigella* spp./EIEC<sup>d</sup> (ipaH) | 2% | 8% | 0% |
| *Plesiomonas shigelloides* | 2% | 8% | 0% |
| *Salmonella* spp. | 2% | 0% | 2% |
| *Yersinia enterocolitica* | 2% | 8% | 0% |
| **Virus** |  |  |  |
| norovirus (GI/II) | 2% | 0% | 2% |
| **Protozoa** |  |  |  |
| *Acanthamoeba* spp. | 12% | 17% | 11% |
| *Cryptosporidium* spp. | 8% | 0% | 11% |
| *Giardia* spp. | 7% | 0% | 9% |
| *Blastocystis* spp. | 3% | 8% | 2% |
| *Balantidium coli* | 2% | 0% | 2% |
| **Soil-transmitted helminth** |  |  |  |
| *Trichuris* spp. | 2% | 0% | 2% |
| <sup>a</sup>Enteropathogenic *Escherichia coli*<br><sup>b</sup>Enteropathogenic *Escherichia coli*<br><sup>c</sup>Enterotoxigenic *Escherichia coli*<br><sup>d</sup>Enteroinvasive *E. coli*<br><sup>e</sup>Classified based on the concentration of human mitochondrial DNA (Supporting Information, Text S5)<sup>19</sup><br>Not detected in any sample: *Campylobacter jejuni/coli*, *Clostridium difficile*, *E. coli* O157:H7, astrovirus, rotavirus, sapovirus (I/II/IV/V), SARS-CoV-2, *Cystoisospora belli*, *Cyclospora cayetanensis*, *Enterocytozoon bienersi*, *Encephalitozoon intestinalis*, *Entamoeba histolytica*, *Entamoeba* spp., *Ancylostoma duodenale*, *Ascaris lumbricoides*, *Enterobius vermicularis*, *Hymenolepis nana*, *Necator americanus*, and *Strongyloides stercoralis* |  |  |  |

The mtDNA dPCR assay indicated 12 samples were likely human in origin and 47 were likely animal in origin (Figure S3). The prevalence of atypical EPEC was higher in human stools (42%) than in the non-human stools (36%). Typical EPEC, tied for second most abundant pathogen found, was absent in human stools, as were *Salmonella*, Norovirus, *Cryptosporidium, Giardia*, and *Balantidium coli*. The pathogens *Helicobacter pylori, Shigella* spp./EIEC (enteroinvasive *Escherichia coli*), *Plesiomonas shigelloides*, and *Yersinia entercolitica* were only found in human fecal samples.

Two samples were TAC-positive for *Trichuris*, one of which was presumptively of human origin. Due to a borderline initial Cq value (34.4), a negative microscopy result, and inhibited or negative results on subsequent TAC runs for *Trichuris* for this sample, we excluded the result in our prevalence calculations. Microscopy for the other (non-human) *Trichuris*-positive sample revealed *Trichuris vulpis, Toxocara canis*, and hookworm ova (Figure S4).

We estimated the annual number of pathogens prevented from release into the local environment as a result of each Pit Stop intervention under three scenarios (Table 2): (i) that fecal waste reduced was both human and animal, since Pit Stop interventions also include animal waste bags and bins (ii) that all fecal waste reduced was of human origin; and (3) (i) that all fecal waste reduced was of animal origin. There results of the three scenarios were similar due to the high concentration of pathogen shedding in feces, except for the instances where we only detected a pathogen in human or non-human feces (e.g., *Cryptosporidium*) (Table 2). The estimated reduction in pathogens released to environment in the scenario that considered both fecal waste sources varied from 1 × 10^7^ (95% CI: 1.6 × 10^6^, 1.3 × 10^8^) for *Trichuris* to 6.3 × 10^12^ (95% CI: 4.0 × 10^12^, 7.9 × 10^12^) for atypical enteropathogenic *E. coli*.

**Table 2.**
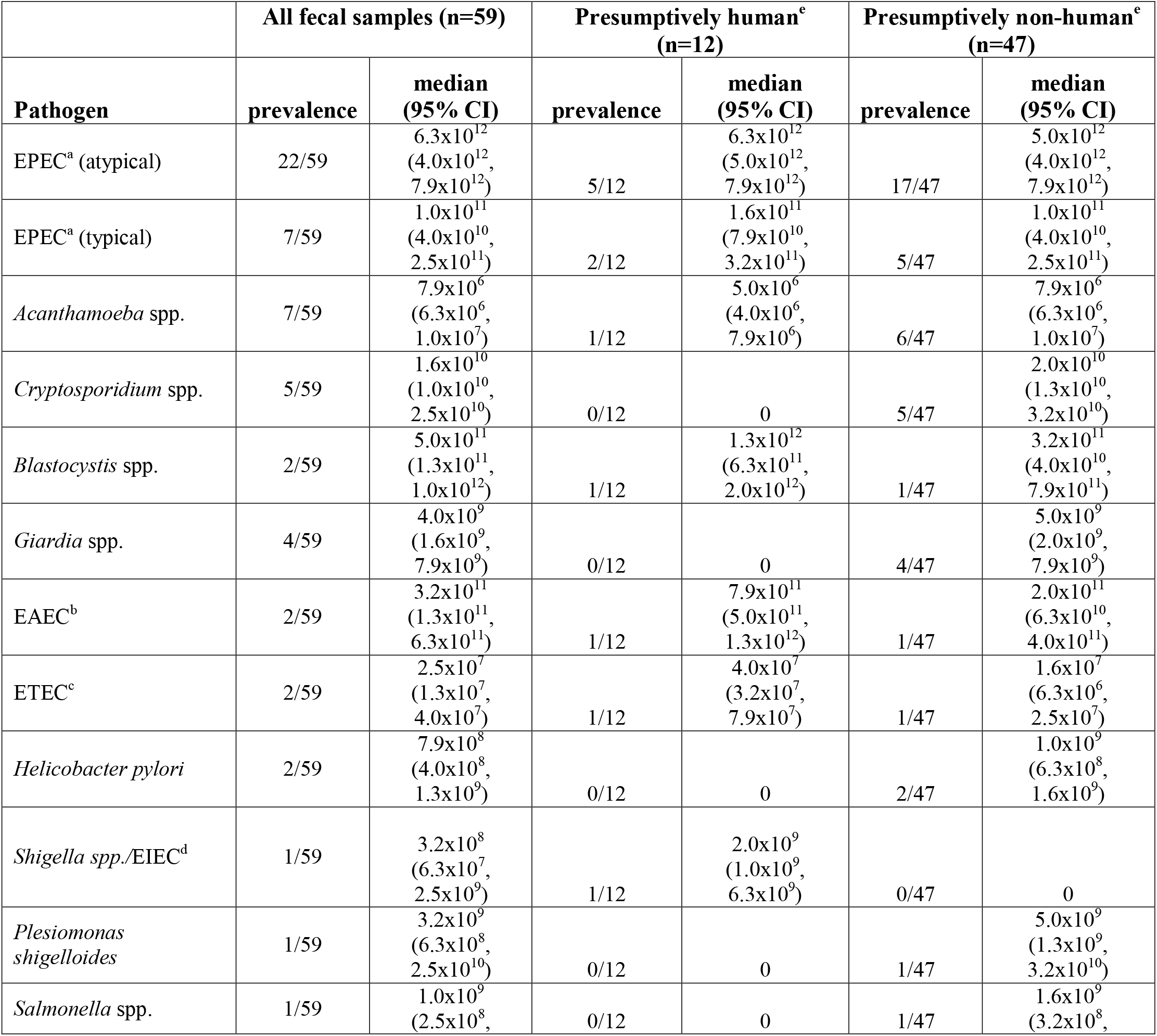
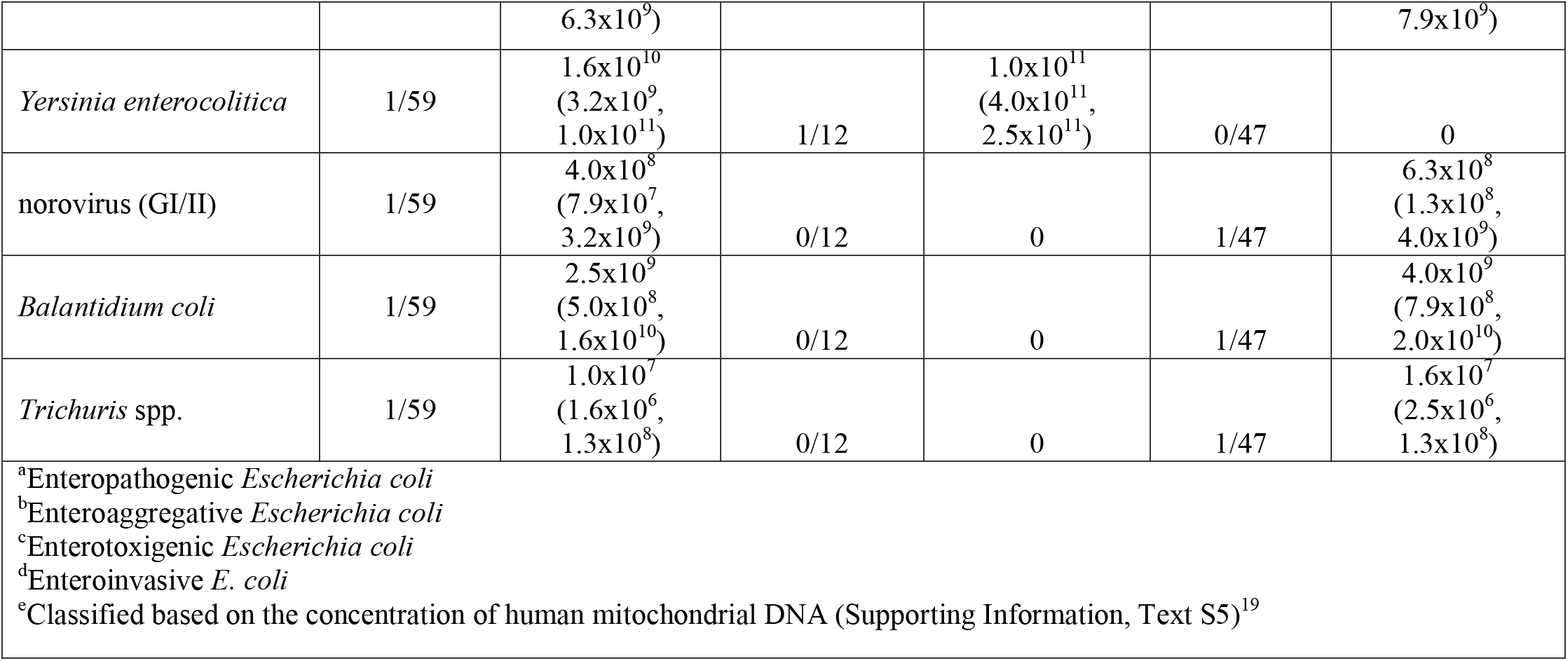
Modeled annual pathogens prevented from release into the environment per Pit Stop in study area (within 500 m walking distance).

|  | All fecal samples (n=59) |  | Presumptively human<sup>e</sup><br>(n=12) |  | Presumptively non-human<sup>e</sup><br>(n=47) |  |
| --- | --- | --- | --- | --- | --- | --- |
| Pathogen | prevalence | median<br>(95% CI) | prevalence | median<br>(95% CI) | prevalence | median<br>(95% CI) |
| EPEC<sup>a</sup> (atypical) | 22/59 | 6.3x10<sup>12</sup><br>(4.0x10<sup>12</sup>,<br>7.9x10<sup>12</sup>) | 5/12 | 6.3x10<sup>12</sup><br>(5.0x10<sup>12</sup>,<br>7.9x10<sup>12</sup>) | 17/47 | 5.0x10<sup>12</sup><br>(4.0x10<sup>12</sup>,<br>7.9x10<sup>12</sup>) |
| EPEC<sup>a</sup> (typical) | 7/59 | 1.0x10<sup>11</sup><br>(4.0x10<sup>10</sup>,<br>2.5x10<sup>11</sup>) | 2/12 | 1.6x10<sup>11</sup><br>(7.9x10<sup>10</sup>,<br>3.2x10<sup>11</sup>) | 5/47 | 1.0x10<sup>11</sup><br>(4.0x10<sup>10</sup>,<br>2.5x10<sup>11</sup>) |
| *Acanthamoeba* spp. | 7/59 | 7.9x10<sup>6</sup><br>(6.3x10<sup>6</sup>,<br>1.0x10<sup>7</sup>) | 1/12 | 5.0x10<sup>6</sup><br>(4.0x10<sup>6</sup>,<br>7.9x10<sup>6</sup>) | 6/47 | 7.9x10<sup>6</sup><br>(6.3x10<sup>6</sup>,<br>1.0x10<sup>7</sup>) |
| *Cryptosporidium* spp. | 5/59 | 1.6x10<sup>10</sup><br>(1.0x10<sup>10</sup>,<br>2.5x10<sup>10</sup>) | 0/12 | 0 | 5/47 | 2.0x10<sup>10</sup><br>(1.3x10<sup>10</sup>,<br>3.2x10<sup>10</sup>) |
| *Blastocystis* spp. | 2/59 | 5.0x10<sup>11</sup><br>(1.3x10<sup>11</sup>,<br>1.0x10<sup>12</sup>) | 1/12 | 1.3x10<sup>12</sup><br>(6.3x10<sup>11</sup>,<br>2.0x10<sup>12</sup>) | 1/47 | 3.2x10<sup>11</sup><br>(4.0x10<sup>10</sup>,<br>7.9x10<sup>11</sup>) |
| *Giardia* spp. | 4/59 | 4.0x10<sup>9</sup><br>(1.6x10<sup>9</sup>,<br>7.9x10<sup>9</sup>) | 0/12 | 0 | 4/47 | 5.0x10<sup>9</sup><br>(2.0x10<sup>9</sup>,<br>7.9x10<sup>9</sup>) |
| EAEC<sup>b</sup> | 2/59 | 3.2x10<sup>11</sup><br>(1.3x10<sup>11</sup>,<br>6.3x10<sup>11</sup>) | 1/12 | 7.9x10<sup>11</sup><br>(5.0x10<sup>11</sup>,<br>1.3x10<sup>12</sup>) | 1/47 | 2.0x10<sup>11</sup><br>(6.3x10<sup>10</sup>,<br>4.0x10<sup>11</sup>) |
| ETEC<sup>c</sup> | 2/59 | 2.5x10<sup>7</sup><br>(1.3x10<sup>7</sup>,<br>4.0x10<sup>7</sup>) | 1/12 | 4.0x10<sup>7</sup><br>(3.2x10<sup>7</sup>,<br>7.9x10<sup>7</sup>) | 1/47 | 1.6x10<sup>7</sup><br>(6.3x10<sup>6</sup>,<br>2.5x10<sup>7</sup>) |
| *Helicobacter pylori* | 2/59 | 7.9x10<sup>8</sup><br>(4.0x10<sup>8</sup>,<br>1.3x10<sup>9</sup>) | 0/12 | 0 | 2/47 | 1.0x10<sup>9</sup><br>(6.3x10<sup>8</sup>,<br>1.6x10<sup>9</sup>) |
| *Shigella* spp./EIEC<sup>d</sup> | 1/59 | 3.2x10<sup>8</sup><br>(6.3x10<sup>7</sup>,<br>2.5x10<sup>9</sup>) | 1/12 | 2.0x10<sup>9</sup><br>(1.0x10<sup>9</sup>,<br>6.3x10<sup>9</sup>) | 0/47 | 0 |
| *Plesiomonas shigelloides* | 1/59 | 3.2x10<sup>9</sup><br>(6.3x10<sup>8</sup>,<br>2.5x10<sup>10</sup>) | 0/12 | 0 | 1/47 | 5.0x10<sup>9</sup><br>(1.3x10<sup>9</sup>,<br>3.2x10<sup>10</sup>) |
| *Salmonella* spp. | 1/59 | 1.0x10<sup>9</sup><br>(2.5x10<sup>8</sup>, | 0/12 | 0 | 1/47 | 1.6x10<sup>9</sup><br>(3.2x10<sup>8</sup>, |
|  |  | 6.3x10<sup>9</sup>) |  |  |  | 7.9x10<sup>9</sup>) |
| *Yersinia enterocolitica* | 1/59 | 1.6x10<sup>10</sup><br>(3.2x10<sup>9</sup>,<br>1.0x10<sup>11</sup>) | 1/12 | 1.0x10<sup>11</sup><br>(4.0x10<sup>11</sup>,<br>2.5x10<sup>11</sup>) | 0/47 | 0 |
| norovirus (GI/II) | 1/59 | 4.0x10<sup>8</sup><br>(7.9x10<sup>7</sup>,<br>3.2x10<sup>9</sup>) | 0/12 | 0 | 1/47 | 6.3x10<sup>8</sup><br>(1.3x10<sup>8</sup>,<br>4.0x10<sup>9</sup>) |
| *Balantidium coli* | 1/59 | 2.5x10<sup>9</sup><br>(5.0x10<sup>8</sup>,<br>1.6x10<sup>10</sup>) | 0/12 | 0 | 1/47 | 4.0x10<sup>9</sup><br>(7.9x10<sup>8</sup>,<br>2.0x10<sup>10</sup>) |
| *Trichuris* spp. | 1/59 | 1.0x10<sup>7</sup><br>(1.6x10<sup>6</sup>,<br>1.3x10<sup>8</sup>) | 0/12 | 0 | 1/47 | 1.6x10<sup>7</sup><br>(2.5x10<sup>6</sup>,<br>1.3x10<sup>8</sup>) |
| <sup>a</sup>Enteropathogenic *Escherichia coli*<br><sup>b</sup>Enteropathogenic *Escherichia coli*<br><sup>c</sup>Enterotoxigenic *Escherichia coli*<br><sup>d</sup>Enteroinvasive *E. coli*<br><sup>e</sup>Classified based on the concentration of human mitochondrial DNA (Supporting Information, Text S5)<sup>19</sup> |  |  |  |  |  |  |

Because the Pit Latrine intervention includes animal waste bags and bins, it is plausible that these facilities may also result in reductions in enteric pathogens observed only in animal feces, including EPEC (typical), *Salmonella* spp., norovirus, *Cryptosporidium* spp., *Giardia* spp., *Balantidium coli*, and *Trichuris* spp. Assuming that the Pit Stop facilities reduce both human and animal feces in the same proportion that they appear in our fecal samples (20% human, 80% non-human), we estimate quantitative reductions of the broad range of pathogens represented across all samples (Table 2).

## DISCUSSION

We detected a wide range of enteric pathogens in the fecal samples we collected, with approximately half (51%) of all samples positive for one or more of the pathogens we sought. Based on our pathogen analysis of fecal wastes and previously estimated reductions in fecal wastes on the street^15^, the Pit Stop intervention has likely reduced the number of pathogens released into the environment within 500 m walking distance of each new toilet facility installed. Reducing pathogen hazards in a densely populated environment can prevent disease transmission, especially for the most vulnerable population: people experiencing homelessness, particularly unsheltered people living in the study area who previously lacked accessible public sanitation and hygiene facilities.

Four out of five samples were non-human in origin, with *Giardia, Cryptosporidium*, and EPEC (typical) occurring only in animal stools, though each of these has zoonotic potential^12, 29^. Microscopy results also revealed a high burden of helminth infection in a presumptive canine sample. Humans are not the definitive host of *Toxocara* and canine hookworm, but humans can be infected by them^30, 31^. While provision of public toilets has the potential to reduce human open defecation, control of animal feces requires different interventions. Although the Pit Stop intervention included animal waste bag distribution and disposal bins, further measures are probably required, including public education, enforcement, environmental controls, or other measures. Stray or feral animals may also contribute fecal waste. Based on our detection of a range of potentially zoonotic enteric pathogens in non-human fecal waste with the potential for human contact, control of animal feces should be considered in this setting^12, 29^.

Our findings should be considered alongside some limitations and caveats. First, though we tested for a range of important enteric pathogens, we selected these targets *a priori* and they are a subset of pathogens that may be relevant in this context, especially considering the widespread presence of non-human fecal wastes. Other potential zoonoses, including *Toxoplasma gondii, Toxocara*, and canine hookworm may have been present and future studies should consider them^12, 29^. Second, the frequency of detection of these pathogens cannot be assumed to represent prevalence of infection in any population: multiple fecal samples in the study area may well have been from a single individual, and our quantitative estimations are based on a limited number of samples. Third, detection of pathogen-associated nucleic acids does not and cannot indicate viability or infectivity. These data cannot be used directly in assessing risk of exposure, without further assumptions beyond the scope of our analysis. Fecal samples were apparently fresh when sampled, but pathogens can be inactivated in the environment. Fourth, even without public toilets, many discarded stools will go on to be collected and safely disposed of through street and sidewalk cleaning, being effectively removed from the environment and therefore unlikely to result in exposure. For this reason and others, we cannot conclude that the reduction in pathogen hazards associated with feces would necessarily result in changes to human exposures, infection, or disease, only that the potential exists.

Fifth, we treated non-detects on TAC as true zeroes, so our estimates might be conservative given the lower limit of detection for targets using these assays (Table S3). While TAC uses the same highly sensitive and specific probe-based RT-qPCR chemistry that has been widely adopted for pathogen detection and quantification in both clinical and research settings across a range of sample matrices, the physical constraints of the platform result in much smaller reaction volumes (∼1.5 μL) than for traditional tube-based approaches (20 μL – 50 μL). The reduced reaction volume may negatively impact analytical sensitivity (i.e., increase the probability of false negatives at low target concentrations) and likewise increase the variability of estimated target quantities at lower concentrations. However, TAC has previously been shown to compare favorably with traditional qPCR approaches in terms of quantification linearity as well as pathogen-specific sensitivity and specificity in stool samples^21, 23, 32^. We observed similar linearity of quantification with our standard curves for all but two assays, which were excluded from the analysis (Table S3), suggesting that our estimated pathogen gene copy concentrations in positive samples were sufficiently reliable for the purposes of parametrizing the log-normal pathogen concentration distributions (which operate on the scale of order-of-magnitude differences in pathogen quantity) we employed in our stochastic models.

Finally, in our quantitative model estimating pathogen reductions attributable to the Pit Stop intervention, we assumed that our stool samples (their pathogen content over time, and human/animal origin) are representative of the fecal wastes reduced due to public toilet construction. This may or may not be the case. Our samples were from a narrow window in time, when some pathogens may be more prevalent than others, and this may not be representative of what is being shed over time in the populations contributing fecal wastes to the streets in our study area. We assumed a mean reduction of 18 instances of OD within 500 meters of newly installed Pit Stop locations^15^ in our study area, per week, throughout the modeled period of one year based on six months of observational data. We first estimated reductions in pathogen hazards assuming all contained waste was of human origin, given the Pit Stop’s primary ostensible role in serving people. We also estimated hazard reductions based on enteric pathogen quantification across all samples, which from our collection effort were determined to be 20% human origin and 80% animal origin. The Pit Stop interventions included both toilet facilities as well as bags and bins for animal waste control, so both types of stools could plausibly be reduced in the immediate surroundings. We observed differences between pathogens detected according to presumptive human versus animal sources, with some pathogens appearing in only human stools and others occurring in non-human stool only. For example, *Helicobacter pylori* and *Shigella*/EIEC were detected only in presumptively human stools.

Clean water, safe sanitation, and adequate hygiene are not universal in American cities^2, 33, 34^, with gaps most apparent among those experiencing homelessness^2, 7^. San Francisco deserves credit for proactively working to solve this problem, which is not inexpensive^14, 15^ and can be politically contentious as cities grapple with the growing crisis of homelessness. Water and sanitation are human rights^35^ that are essential to living a dignified life. Construction of publicly accessible, safe toilets is a commonsense approach to reducing enteric pathogen hazards in cities^9^, though the primary reason to continue to invest in public sanitation facilities is to support the physical, mental, and social well-being of people – much of which is difficult or impossible to measure in practice – and because it is the right and humane thing to do. Moreover, sanitation is a biological necessity that is needed wherever people live. The waste must go somewhere: an adult weighing 80 kg (near the mean body mass in North America) will produce an average of approximately 38 kg of feces per year^36, 37^, 100% of which should be effectively and safely managed to protect all members of the community from infectious disease. Cities should also consider interventions aimed at reducing animal feces, which are an underappreciated source of enteric pathogen hazards in urban spaces. Our findings demonstrate that a wide range of pathogens with zoonotic potential may be present in discarded animal waste, with uncertain implications for human exposure and disease transmission.

## Data Availability

All data produced in the present study are available upon reasonable request to the authors

## Notes

### Competing Interest Statement

The authors have declared no competing interest.

### Funding Statement

This study did not receive any funding.

### Summary of Updates

The order of the author list is incorrect. I have updated the author list to match the manuscript we have submitted. The change is to change the last author to Joe Brown.

