## Supporting Information for "Public toilets have reduced enteric pathogen hazards in San Francisco"

**Summary: Supporting information contains 4 text additions, 7 tables, and 4 figures, plus references**

Contents

1. Text S1. Ground-truthing 311 data on fecal waste
2. Text S2. Stool pre-treatment protocol
3. Text S3. TAC protocol
4. Text S4. Mini-FLOTAC ova enumeration procedure
5. Text S5. dPCR assay
6. Text S6. Positive and negative control results
7. Table S1. Human mtDNA dPCR assay primer and probe sequences
8. Table S2. TAC primer and probe sequences for pathogen detection
9. Table S3. TAC performance and standard curves
10. Table S4. MIQE Checklist
11. Table S5. Quantitative model parameters
12. Table S6**.** Quantal detection data
13. Figure S1. Intervention and sample location, central San Francisco
14. Figure S2. Intervention toilet
15. Figure S3. Plots of human mtDNA concentration by sample
16. Figure S4. Microscopy images
17. Figure S5. mtDNA dPCR 2-D scatterplots
18. Figure S6. qPCR amplification and multicomponent plots
19. Table S7. dMIQE checklist for mtDNA dPCR
20. Supporting References

**Text S1.** Ground-truthing 311 data on fecal waste.

Methods. The available 311 data include any reports of fecal waste in the city of San Francisco, including both presumed animal or human feces (they are not distinguished separately in the reporting system). Because we sought to use 311 reports of fecal waste to estimate feces-associated pathogen reductions attributable to the Pit Stop facilities, we conducted a limited ground survey of an area in the project area with spatially and temporally matched 311 data. In brief, we chose a weekday and a weekend day to walk a pre-defined route in an area with a high number of 311 reports, including Ellis, Leavenworth, Jones, and O’Farrell streets in the Tenderloin District. We collected data on any fecal waste observed, including both presumed animal or human waste. We timed these surveys before street cleaning so we could match 311 data for these same areas with our direct observations.

Results. We observed 31 instances of fecal waste along our pre-defined route. Available data from the 311 system recorded 25 instances over the same area in the previous 24 hours, so we concluded that 311 data may accurately, but possibly conservatively, reflect fecal waste in the study area.

**Text S2.** Stool pre-treatment protocol.

We added 100 mg of stool into Precellys® SK38 bead beating tubes (Bertin Corp, Rockville, MD) containing 1 mL of Qiagen Buffer ASL (Qiagen, Hilden, Germany) using inoculating loops. We spiked in 10^6^ copies of a gBlock (IDT, Coralville, IA) matching sequence for phocine herpesvirus target used as a DNA extraction positive control and spiked in 10^7^ copies of phage MS2 as an RNA extraction positive control. We vortexed the bead beating tubes for 5 minutes, incubated them at room temperature for 15 minutes, and then centrifuged them at 14,000 rpm for 2 minutes. We proceeded with extraction (eluent volume 100 µL) following the manufacturer’s protocol for the QIAamp 96 Virus QIAcube HT Kit, which we automated on the QIAcube (Qiagen, Hilden, Germany).

**Text S3.** Taqman Array Card (TAC) protocol.

We immediately stored extracted nucleic acids at 4 °C for no more than 24 hours before transferring to 0.6 mL centrifuge tubes and storing at -80 °C.

We developed an engineered combined positive control for all the targets on our custom TAC following the approach of Kodani and Winchell (2012)^1^, as described previously^2^. Briefly, the primers and probe sequences for all targets were concatenated and synthesized on two plasmids (GeneArt, Regensburg, Germany), one containing the DNA targets and the other the RNA targets. The DNA-target plasmid used directly while the RNA-target plasmid was linearized with a BshT1 restriction enzyme (Thermofisher, Waltham, MA) and transcribed (MEGAscript T7 Transcription Kit and MEGAclear Transcription Clean-Up Kit, Thermofisher, Waltham, MA) to generate RNA control material. The DNA plasmid concentration (copies/µL) was determined from the manufacturer-provided mass and the transcribed RNA plasmid was quantified using a Qubit RNA HS Assay Kit on Qubit 4 Fluorometer (Thermofisher, Waltham, MA). The two plasmids were mixed 1:1 by concentration, aliquoted in 0.6 mL centrifuge tubes, and stored at -80 °C for single use to minimize freeze-thaw cycles. We used the combined plasmids at stock concentration on one TAC card per day of analysis as a positive control and also as standard reference material to generate standard curves for target quantification.

For analysis, we mixed 40 μL of total nucleic acid template (0.6 μL template per reaction well) with 60 μL master mix composed of AgPath-ID™ One-Step RT-PCR Reagents (Thermo Fisher Scientific, Waltham, MA), then loaded each port with a combined 100 μL. For the first card on a given day, one port was run with a PCR positive control and one with a PCR negative control (i.e., molecular water).

Following manufacturer instructions, we centrifuged each card twice at 1200 rpm for one minute, sealed the card, trimmed the loading ports, and loaded the card into a QuantStudio 7 (Thermo Fisher Scientific, Waltham, MA). To perform quantitative reverse transcription real-time PCR, we used the following cycling conditions with a 1 °C/s ramp rate between all steps: 45 °C for 20 minutes, 95 °C for 10 minutes, then 45 cycles of 95 °C for 15 seconds and 60 °C for 1 minute.

TAC performance was evaluated using an 8-fold dilution series (10^9^-10^2^ gene copies per reaction well) of the engineered combined positive control. The linearity and efficiency for 36 of the 38 targets were within normative standards (linearity: 0.97-1.0, efficiency: 87%-102%). The assays for hepatitis A virus and adenovirus 40/41 did not perform well and were excluded from analysis.

**Text S4.** Microscopy: ova enumeration procedure.

The Mini-FLOTAC system^3^ used to separate ova from stool, using 625-1050 mg of wet weight stool and 36 mL of 1.25 specific gravity sodium nitrate solution to float ova in 3 cartridges with 2 aliquots on each cartridge. Cartridges were then read on a grid at 100x magnification, and any ova were speciated (if possible) and counted^4^.

**Text S5.** dPCR assay.

The hCYTB484 assay (Table S1) has previously demonstrated a nearly 100% sensitivity and specificity for detecting human mitochondrial DNA across multiple populations and geographies^5^. We performed dPCR reactions containing 2 µL of template (without added restriction enzyme), 800 nM final concentration for both forward and reverse primers, 400 nM Sun Probe (IDT, Coralville, IA), and 4X QIAcuity PCR MasterMix, with nuclease-free water added to achieve a final reaction volume of 40 µL. Cycling conditions consisted of 2 minutes at 95°C, followed by 40 cycles of 15s at 95°C and 30s at 59°C. The reaction mixture was loaded onto QIAcuity Nanoplate 26k 24-well plates and run on a QIAGEN QIAcuity Four machine operated using the QIAcuity Software Suite 2.1.7.182 (Qiagen, Hilden, Germany). Positive PCR controls (samples of known human origin, n = 4) were run in addition to negative PCR controls (no template controls, n = 6). Additionally, we analyzed technical replicates for 25% of samples (n = 15), for which qualitative results (i.e. positive/negative) were consistent, demonstrating repeatability. Clear separation between bands of positive and negative partitions, (or difference in fluorescence amplitude between negative and positive partitions), respectively, was observed for both positive PCR controls and experimental samples (Figure S5). Thresholds (partition classification) were either automatically or manually set above the negative partitions for each plate run. to differentiate between negative and positive partitions according to their fluorescence (RFUs). Please see Figure S5 for images of dPCR RFU plots. The average number of partitions per reaction was 24879 (standard deviation = 1590). Full details on the development of the assay, including methods used for design, in silico verification, optimization, and location of amplicon, can be found in Zhu *et al*., 2020^5^.

**Table S1**. hCYTB484 human mtDNA primers and probe sequences.

| Oligonucleotide | Sequence (5’ to 3’) | Reference | Notes |
| --- | --- | --- | --- |
| hCYTB484F | CAATGAATCTGAGGAGGCTAC | Zhu *et al*. 2020^5^ | 121 bp amplicon |
| hCYTB604R | CGTGCAAGAATAGGAGGTG | Zhu *et al*. 2020^5^ |  |
| hCYTB520TM | FAM-ACCCTCACACGATTCTTTACCTTTCACT-BHQ | Zhu *et al*. 2020^5^ |  |

**Table S2.** Primer and probe sequences for qPCR assays on the custom TAC

| **Pathogen** | **Gene** | **Primer or probe sequence (5' - 3')** | **Reference** |
| --- | --- | --- | --- |
| Astrovirus | Capsid | Fwd: CAGTTGCTTGCTGCGTTCA | Liu *et al*. 2016^2^ |
|  |  | Rev: CTTGCTAGCCATCACACTTCT |  |
|  |  | Probe: CACAGAAGAGCAACTCCATCGC |  |
| Pan-enterovirus | 5'UTR | Fwd: CCCTGAATGCGGCTAATCC | Liu *et al*. 2016^2^ |
|  |  | Rev: GCGATTGTCACCATWAGCAG |  |
|  |  | Probe: CCGACTACTTTGGGWGTCCGT |  |
| Norovirus GI | ORF1-ORF2 | Fwd: CGYTGGATGCGNTTYCATGA | Liu *et al*. 2016^2^ |
|  |  | Rev: CTTAGACGCCATCATCATTYAC |  |
|  |  | Probe: TGGACAGGAGATCGC |  |
| Norovirus GII | ORF1-ORF2 | Fwd: CARGARBCNATGTTYAGRTGGATGAG | Liu *et al*. 2016^2^ |
|  |  | Rev: TCGACGCCATCTTCATTCACA |  |
|  |  | Probe: TGGGAGGGCGATCGCAATCT |  |
| Sapovirus (I, II, IV) | RdRp | Fwd: GAYCAGGCTCTCGCYACCTAC | Liu *et al*. 2016^2^ |
|  |  | Rev: CCCTCCATYTCAAACACTA |  |
|  |  | Probe: CYTGGTTCATAGGTGGTRCAG |  |
| Sapovirus V | RdRp | Fwd: TTTGAACAAGCTGTGGCATGCTAC | Liu *et al*. 2016^2^ |
|  |  | Rev: CCCTCCATYTCAAACACTA |  |
|  |  | Probe: CAGCTGGTACATTGGTGGCAC |  |
| Adenovirus 40/41 | Hexon b | Fwd: AACTTTCTCTCTTAATAGACGCC | Liu *et al*. 2016^2^ |
|  |  | Rev: AGGGGGCTAGAAAACAAAA |  |
|  |  | Probe: CTGACACGGGCACTCT |  |
| Rotavirus | NSP3 | Fwd: ACCATCTWCACRTRACCCTCTATGAG | Liu *et al*. 2016^2^ |
|  |  | Rev: GGTCACATAACGCCCCTATAGC |  |
|  |  | Probe: AGTTAAAAGCTAACACTGTCAAA |  |
| *Campylobacter jejuni*/*C. coli* | cadF | Fwd: CTGCTAAACCATAGAAATAAAATTTCTCAC | Liu *et al*. 2016^2^ |
|  |  | Rev: CTTTGAAGGTAATTTAGATATGGATAATCG |  |
|  |  | Probe: CATTTTGACGATTTTTGGCTTGA |  |
| *C. difficile* | tcdB | Fwd: GGTATTACCTAATGCTCCAAATAG | Liu *et al*. 2016^2^ |
|  |  | Rev: TTTGTGCCATCATTTTCTAAGC |  |
|  |  | Probe: CCTGGTGTCCATCCTGTTTC |  |
| EAEC (aaiC) | *aaiC* | Fwd: ATTGTCCTCAGGCATTTCAC | Liu *et al*. 2016^2^ |
|  |  | Rev: ACGACACCCCTGATAAACAA |  |
|  |  | Probe: TAGTGCATACTCATCATTTAAG |  |
| EAEC (aatA) | *aatA* | Fwd: CTGGCGAAAGACTGTATCAT | Liu *et al*. 2016^2^ |
|  |  | Rev: TTTTGCTTCATAAGCCGATAGA |  |
|  |  | Probe: TGGTTCTCATCTATTACAGACAGC |  |
| STEC (stx1) | *stx1* | Fwd: ACTTCTCGACTGCAAAGACGTATG | Liu *et al*. 2016^2^ |
|  |  | Rev: ACAAATTATCCCCTGWGCCACTATC |  |
|  |  | Probe: 56FAM/CTCTGCAATAGGTACTCCA/3MGB-NFQ/ |  |
| STEC (stx2) | *stx2* | Fwd: CCACATCGGTGTCTGTTATTAACC | Liu *et al*. 2016^2^ |
|  |  | Rev: GGTCAAAACGCGCCTGATAG |  |
|  |  | Probe: 5VIC/TTGCTGTGGATATACGAGG/3MGB-NFQ/ |  |
| EPEC (eae) | *eae* | Fwd: CATTGATCAGGATTTTTCTGGTGATA | Liu *et al*. 2016^2^ |
|  |  | Rev: CTCATGCGGAAATAGCCGTTA |  |
|  |  | Probe: 56FAM/ATACTGGCGAGACTATTTCAA/3MGB-NFQ/ |  |
| EPEC (bfpA) | *bfpA* | Fwd: TGGTGCTTGCGCTTGCT | Liu *et al*. 2016^2^ |
|  |  | Rev: CGTTGCGCTCATTACTTCTG |  |
|  |  | Probe: 5VIC/CAGTCTGCGTCTGATTCCAA/3MGB-NFQ/ |  |
| ETEC LT | *LT* | Fwd: TTCCCACCGGATCACCAA | Liu *et al*. 2016^2^ |
|  |  | Rev: CAACCTTGTGGTGCATGATGA |  |
|  |  | Probe: CTTGGAGAGAAGAACCCT |  |
| ETEC ST | *ST* | Fh, GCTAAACCAGYAGRGTCTTCAAAA | Liu *et al*. 2016^2^ |
|  |  | Fp, TGAATCACTTGACTCTTCAAAA |  |
|  |  | Rh, CCCGGTACARGCAGGATTACAACA |  |
|  |  | Rp, GGCAGGATTACAACAAAGTT |  |
|  |  | Ph, 6VIC/TGGTCCTGAAAGCATGAA/3MGB-NFQ/ |  |
|  |  | Pp, 6VIC/TGAACAACACATTTTACTGCT/3MGB-NFQ/ |  |
| EIEC/*Shigella* | *ipaH* | Fwd: CCTTTTCCGCGTTCCTTGA | Liu *et al*. 2016^2^ |
|  |  | Rev: CGGAATCCGGAGGTATTGC |  |
|  |  | Probe: 56FAM/CGCCTTTCCGATACCGTCTCTGCA/3MGB-NFQ/ |  |
| *Salmonella* | *ttr*  MGB probe | Fwd: CTCACCAGGAGATTACAACATGG | Liu *et al*. 2016^2^ |
|  |  | Rev: AGCTCAGACCAAAAGTGACCATC |  |
|  |  | Probe: CACCGACGGCGAGACCGACTTT |  |
| *E. coli* O157 | *rfbE* | Fwd: TTTCACACTTATTGGATGGTCTCAA | Liu *et al*. 2016^2^ |
|  |  | Rev: CGATGAGTTTATCTGCAAGGTGAT |  |
|  |  | Probe: CTCTCTTTCCTCTGCGGTCCT |  |
| *Cryptosporidium*  (pan-*Crypto*) | *18S* | Fwd: GGGTTGTATTTATTAGATAAAGAACCA | Liu *et al*. 2016^2^ |
|  |  | Rev: AGGCCAATACCCTACCGTCT |  |
|  |  | Probe: TGACATATCATTCAAGTTTCTGAC |  |
| *Giardia* spp. | *18S* | Fwd: GACGGCTCAGGACAACGGTT | Liu *et al*. 2016^2^ |
|  |  | Rev: TTGCCAGCGGTGTCCG |  |
|  |  | Probe: CCCGCGGCGGTCCCTGCTAG |  |
| *E. histolytica* | *18S* | Fwd: ATTGTCGTGGCATCCTAACTCA | Liu *et al*. 2016^2^ |
|  |  | Rev: GCGGACGGCTCATTATAACA |  |
|  |  | Probe: TCATTGAATGAATTGGCCATTT |  |
| *Entamoeba* spp. | *18S rRNA* | Fwd: AAACGATGTCAACCAAGGATTG | Liu *et al*. 2016^2^ |
|  |  | Rev: TCCCCCTGAAGTCCATAAACTC |  |
|  |  | Probe: CCTTGTTCAGAACTTAAAGAGAAA |  |
| *Ascaris* | *ITS1* | Fwd: GCCACATAGTAAATTGCACACAAAT | Liu *et al*. 2016^2^ |
|  |  | Rev: GCCTTTCTAACAAGCCCAACAT |  |
|  |  | Probe: TTGGCGGACAATTGCATGCGAT |  |
| *Trichuris* | *18S rRNA* | Fwd: TTGAAACGACTTGCTCATCAACTT | Liu *et al*. 2016^2^ |
|  |  | Rev: CTGATTCTCCGTTAACCGTTGTC |  |
|  |  | Probe: CGATGGTACGCTACGTGCTTACCATGG |  |
| *Necator americanus* | ITS-2 | Fwd: CTGTTTGTCGAACGGTACTTGC | Liu *et al*. 2016^2^ |
|  |  | Rev: ATAACAGCGTGCACATGTTGC |  |
|  |  | Probe: CTGTACTACGCATTGTATAC |  |
| *Strongyloides stercoralis* | dispersed repetitive sequence | Fwd: TCCAGAAAAGTCTTCACTCTCCAG | Liu *et al*. 2016^2^ |
|  |  | Rev: TGCGTTAGAATTTAGATATTATTGTTGCT |  |
|  |  | Probe: TCAGCTCCAGTTGAACAACAGCCTCCAA |  |
| *Blastocystis* spp. | 18s rRNA | Fwd: TGGTCCGRTGAACACTTTGGAT | Liu *et al*. 2016^2^ |
|  |  | Rev: CCTACGGAAACCTTGTTACGACTTCA |  |
|  |  | Probe: CTTCCTCTAAATGRTAAGATT |  |
| *Ancylostoma duodenales* | ITS-2 | Fwd: GAATGACAGCAAACTCGTTGTTG | Liu *et al*. 2016^2^ |
|  |  | Rev: ATACTAGCCACTGCCGAAACGT |  |
|  |  | Probe: ATCGTTTACCGACTTTAG |  |
| *Enterobius vermicularis* | 5S rRNA | Fwd: CAAACAACTGCATCACCAATAAC | Rudko et al. 2017^6^ |
|  |  | Rev: AGTGTAGAGCAATAAGCAGTAAAG |  |
|  |  | Probe: TACCAACAACACTTGCACGTCTCTTCA |  |
| *Hymenolepis nana* | ITS1 | Fwd: CATTGTGTACCAAATTGATGATGAGTA |  |
|  |  | Rev: CAACTGACAGCATGTTTCGATATG | Kann *et al*. 2022^7^ |
|  |  | Probe: CGTGTGCGCCTCTGGCTTACCG |  |
| SARS-CoV-2 | N2 | Fwd: TTACAAACATTGGCCGCAAA |  |
|  |  | Rev: GCGCGACATTCCGAAGAA | Lu *et al*. 2020^8^ |
|  |  | Probe: ACAATTTGCCCCCAGCGCTTCAG |  |
| 16S ribosomal RNA | 16S | Fwd: TGCAAGTCGAACGAAGCACTTTA | Liu *et al*. 2016^2^ |
|  |  | Rev: GCAGGTTACCCACGCGTTAC |  |
|  |  | Probe: CGCCACTCAGTCACAAA |  |
| Phocine herpesvirus (PhHV) | gB | Fwd: GGGCGAATCACAGATTGAATC | Liu *et al*. 2016^2^ |
|  |  | Rev: GCGGTTCCAAACGTACCAA |  |
|  |  | Probe: TATGTGTCCGCCACCATCT |  |
| *Yersinia enterocolitica* | *lytA* | Fwd: TGATTCACCAGCAGCAATAC | Liu *et al*. 2016^2^ |
|  |  | Rev: GGCATCATGAAAGGCGG |  |
|  |  | Probe: TGTCGGTTTCTCCTTCCAGG |  |
| *Heliobacter pylori* | *ureC* | Fwd: GACACCAGAAAAAGCGGCTA | Liu *et al*. 2016^2^ |
|  |  | Rev: AGCGCATGTCTTCGGTTAAA |  |
|  |  | Probe: TCACTAAAGCGTTTTCTACC |  |
| *Plesiomonas shigelloides* | *gyrB* | Fwd: CCGCCGTGAAGGCAAAG | Liu *et al*. 2016^2^ |
|  |  | Rev: GCTACCGGCTCACCCAGAT |  |
|  |  | Probe: CACACCCAAGAATAC |  |
| *Cyclospora cayetanensi* | 18s rRNA | Fwd: AAAAGCTCGTAGTTGGATTTCTG | Liu *et al*. 2016^2^ |
|  |  | Rev: AACACCAACGCACGCAGC |  |
|  |  | Probe: AAGGCCGGATGACCACGA |  |
| *Cystoisospora belli* | 18s rRNA | Fwd: ATATTCCCTGCAGCATGTCTGTTT | Liu *et al*. 2016^2^ |
|  |  | Rev: CCACACGCGTATTCCAGAGA |  |
|  |  | Probe: CAAGTTCTGCTCACGCGCTTCTGG |  |
| *Blastocystis* spp. | 18s rRNA | Fwd: TGGTCCGRTGAACACTTTGGAT | Liu *et al*. 2016^2^ |
|  |  | Rev: CCTACGGAAACCTTGTTACGACTTCA |  |
|  |  | Probe: CTTCCTCTAAATGRTAAGATT |  |
| *Enterocytozoon bieneusi* | SSU rRNA | Fwd: TGTGTAGGCGTGAGAGTGTATCTG | Liu *et al*. 2016^2^ |
|  |  | Rev: CATCCAACCATCACGTACCAATC |  |
|  |  | Probe: CACTGCACCCACATCCCTCACCCTT |  |
| *Encephalitozoon intestinalis* | ITS | Fwd: CACCAGGTTGATTCTGCCTGAC | Liu *et al*. 2016^2^ |
|  |  | Rev: CTAGTTAGGCCATTACCCTAACTACCA |  |
|  |  | Probe: CTATCACTGAGCCGTCC |  |
| *Balantidium coli* | ITS-1 | Fwd: TGCAATGTGAATTGCAGAACC | Sow *et al*. 2017^9^ |
|  |  | Rev: TGGTTACGCACACTGAAACAA |  |
|  |  | Probe: CTGGTTTAGCCAGTGCCAGTTGC |  |
| *Acanthamoeba* spp. | 18S rRNA | Fwd: CCCAGATCGTTTACCGTGAA | Qvarnstrom *et al*. 2006^10^ |
|  |  | Rev: TAAATATTAATGCCCCCAACTATC |  |
|  |  | Probe: CTGCCACCGAATACATTAGCATGG |  |
| Hepatitis A Virus | 5' NCR | Fwd: TCACCGCCGTTTGCCTAG | Costafreda et al. 2006^11^ |
|  |  | Rev: GGAGAGCCCTGGAAGAAAG |  |
|  |  | Probe: TTAATTCCTGCAGGTTCAGG |  |
| SARS-CoV-2 | N1 | Fwd: GACCCCAAAATCAGCGAAAT | Lu *et al*. 2020^8^ |
|  |  | Rev: TCTGGTTACTGCCAGTTGAATCTG |  |
|  |  | Probe: ACCCCGCATTACGTTTGGTGGACC |  |

**Table S3.** TaqMan Array Card (TAC) performance and standard curve parameters**.**

| **Target** | **Target Gene** | **Slope** | **Y-intercept** | **R^2^** | **Efficiency** | **95% LOD†** |
| --- | --- | --- | --- | --- | --- | --- |
| enteric 16S | 16S | -3.309 | 38.881 | 0.998 | 101% | 0.6 |
| *Acanthamoeba* spp. | 18S rRNA | -3.3877 | 37.82 | 1.000 | 97% | 23 |
| Adenovirus 40/41* | Fiber gene | NA | NA | 0.670 | NA | NA |
| *Ancylostoma duodenale* | ITS-2 | -3.3832 | 39.101 | 1.000 | 98% | 6.2 |
| *Ascaris lumbricoides* | ITS-1 | -3.4482 | 38.594 | 1.000 | 95% | 6.2 |
| astrovirus |  | -3.6867 | 37.531 | 0.998 | 87% | 6.2 |
| *Balantidium coli* | ITS-1 | -3.3935 | 37.92 | 1.000 | 97% | 2.2 |
| *Blastocystis* spp. | 18S rRNA | -3.3196 | 40.64 | 0.997 | 100% | 2.2 |
| *Cystoisospora belli* | 18S rRNA | -3.3479 | 37.801 | 0.999 | 99% | 6.2 |
| *Cyclospora cayetanensi* | 18S rRNA | -3.3408 | 37.151 | 0.998 | 99% | 2.2 |
| *Campylobacter jejuni/coli* | *cadF* | -3.34178 | 38.27 | 0.999 | 99% | 21 |
| *Clostridium difficile* | *tcdB* | -3.4282 | 37.542 | 0.999 | 96% | 6.2 |
| *Cryptosporidium* spp. | 18S rRNA | -3.4033 | 37.983 | 0.999 | 97% | 0.6 |
| DNA control (phocine herpes vírus) | *gB* | -3.315 | 37.009 | 0.998 | 100% | 6.2 |
| *Enterocytozoon bieneusi* | ITS | -3.2802 | 37.209 | 0.999 | 102% | 4.8 |
| *E. coli* O157:H7 | *rfbE* | -3.4568 | 37.976 | 1.000 | 95% | 2.2 |
| *Encephalitozoon intestinalis* | SSU rRNA | -3.3819 | 38.462 | 0.999 | 98% | 2.2 |
| *Enterobius vermicularis* | 5S | -3.4592 | 38.572 | 0.999 | 95% | 72 |
| EAEC (aaiC) | *aaiC* | -3.4241 | 38.15 | 0.999 | 96% | 6.2 |
| EAEC (aatA) | *aatA* | -3.4252 | 37.694 | 0.998 | 96% | 23 |
| *Entamoeba hystolytica* | 18S rRNA | -3.2775 | 37.994 | 0.996 | 102% | 6.2 |
| *Entamoeba* spp. | 18S rRNA | -3.2317 | 37.259 | 0.974 | 104% | 21 |
| EPEC (typical) | *bfpA* | -3.3772 | 37.465 | 0.999 | 98% | 6.2 |
| EPEC (atypical) | *eae* | -3.372 | 37.592 | 0.999 | 98% | 2.2 |
| ETEC (LT) | *LT* | -3.4638 | 47.637 | 0.990 | 94% | 291 |
| ETEC (STh) | *STh* | -3.3785 | 38.763 | 0.999 | 98% | 6.2 |
| ETEC (STp) | *STp* | -3.3548 | 37.266 | 0.999 | 99% | 2.2 |
| *Giardia* spp. | 18S rRNA | -3.4182 | 37.863 | 1.000 | 96% | 6.2 |
| *Hymenolepis nana* |  | -3.3804 | 38.248 | 1.000 | 98% | 2.2 |
| *Helicobacter pylori* | *ureC* | -3.4078 | 37.726 | 0.998 | 97% | 6.2 |
| hepatitis A virus* | NCR | -2.7348 | NA | 0.840 | 132% | NA |
| *Shigella*/EIEC | *ipaH* | -3.3522 | 37.506 | 0.999 | 99% | 23 |
| MS2 | *MS2g1* | -3.5809 | 37.537 | 0.999 | 90% | 1.0 |
| *Necator americanus* | ITS-2 | -3.3686 | 39.806 | 1.000 | 98% | 4.3 |
| Norovirus GII | ORF1-2 | -3.5361 | 36.954 | 0.999 | 92% | 23 |
| Norovirus GI | ORF1-2 | -3.4931 | 35.915 | 0.997 | 93% | 23 |
| *Plesiomonas shigelloides* | *gyrB* | -3.4184 | 38.202 | 1.000 | 96% | 23 |
| rotavirus | NSP3 | -3.5696 | 38.016 | 0.998 | 91% | 6.2 |
| *Salmonella* spp. | *invA* | -3.4191 | 38.427 | 1.000 | 96% | 2.2 |
| Sapovirus I/II/IV | RdRp | -3.6629 | 38.155 | 0.998 | 88% | 2.2 |
| Sapovirus V | RdRp | -3.5616 | 36.69 | 0.999 | 91% | 2.2 |
| SARS-CoV-2 | N1 | -3.5311 | 36.184 | 0.995 | 92% | 6.2 |
| *Strongyloides stercolaris* | Dispersed repetitive sequence | -3.3316 | 37.527 | 0.999 | 100% | 2.2 |
| STEC (stx1) | *stx1* | -3.4083 | 39.883 | 1.000 | 97% | 72 |
| STEC (stx2) | *stx2* | -3.3697 | 38.328 | 0.967 | 98% | 96 |
| *Trichuris trichiura* | 18S rRNA | -3.3502 | 38.395 | 1.000 | 99% | 2.2 |
| *Yersinia enterocolitica* | *lytA* | -3.483 | 38.279 | 0.998 | 94% | 2.2 |

*Excluded due to poor standard curve performance
†95% LOD in gene copies per reaction calculated using methods from Stokdyk *et al*. 2016^12^

**Table S4**. MIQE Checklist

| **ITEM TO CHECK** | **IMPORTANCE** | **CHECKLIST** |
| --- | --- | --- |
| **EXPERIMENTAL DESIGN** |  |  |
| Definition of experimental and control groups | **E** | Cross-sectional study with no intervention or control group |
| Number within each group | **E** | 60 discarded stools were analyzed |
| Assay carried out by core lab or investigator's lab? | D | Investigator's lab |
| **SAMPLE** |  |  |
| Description | **E** | 100 mg stool (see Methods section) |
| Volume/mass of sample processed | D | 100 mg |
| Microdissection or macrodissection | **E** | Not applicable |
| Processing procedure | **E** | Frozen at -80C and shipped on dry ice |
| If frozen - how and how quickly? | **E** | Same day as collection |
| If fixed - with what, how quickly? | **E** | Not fixed |
| Sample storage conditions and duration (especially for FFPE samples) | **E** | N/A |
| **NUCLEIC ACID EXTRACTION** |  |  |
| Procedure and/or instrumentation | **E** | See methods section |
| Name of kit and details of any modifications | **E** | QIAamp 96 Virus QIAcube HT Kit automated on a QIAcube HT |
| Source of additional reagents used | D | Precellys® SK38 bead beating tubes (Bertin Technologies, Rockville, MD) |
| Details of DNase or RNAse treatment | **E** | Not applicable |
| Contamination assessment (DNA or RNA) | **E** | At least one extraction negative control was included during each day of extractions |
| Nucleic acid quantification | **E** | Qubit 1X HS dsDNA Kit |
| Instrument and method | **E** | Qubit 4 Fluorometer |
| RNA integrity method/instrument | **E** | Not measured |
| Inhibition testing (Cq dilutions, spike or other) | **E** | Monitored amplification of spiked controls |
| **REVERSE TRANSCRIPTION** |  |  |
| Complete reaction conditions | **E** | One-step reverse transcription |
| Amount of RNA and reaction volume | **E** | Reaction Volume = 1.5 µL |
| Priming oligonucleotide (if using GSP) and concentration | **E** | Proprietary |
| Reverse transcriptase and concentration | **E** | ArrayScript™ Reverse Transcriptase |
| Temperature and time | **E** | 45°C for 20 minutes |
| Manufacturer of reagents and catalogue numbers | D | Applied Biosystems, AgPath-ID™ One-Step RT-PCR Reagents, Catalog number: 4387391 |
| **qPCR TARGET INFORMATION** |  |  |
| If multiplex, efficiency and LOD of each assay. | **E** | Table S3 |
| *In silico* specificity screen (BLAST, etc) | **E** | We BLASTed all assays to confirm specificity before ordering the custom TAC. |
| **qPCR OLIGONUCLEOTIDES** |  |  |
| Primer sequences | **E** | Table S2 |
| Probe sequences | D** | Table S2 |
| Location and identity of any modifications | **E** | No modifications |
| Manufacturer of oligonucleotides | D | ThermoFisher Scientific |
| **qPCR PROTOCOL** |  |  |
| Complete reaction conditions | **E** | 45°C for 20 min and 95°C for 10 min, followed by 45 cycles of 95°C for 15 s and 60°C for 1 min |
| Reaction volume and amount of cDNA/DNA | **E** | 40 µL of template with 60 µL of AgPath-ID™ One-Step RT-PCR Reagents |
| Primer, (probe), Mg++ and dNTP concentrations | **E** | All assays contained the same concentrations of primers (900 nanomolar) and probe (250 nanomolar). The Mg2+ and dNTP concentrations are not listed in the in the User Guide. |
| Polymerase identity and concentration | **E** | AmpliTaq Gold™ polymerase |
| Buffer/kit identity and manufacturer | **E** | AgPath-ID™ One-Step RT-PCR Reagents |
| Additives (SYBR Green I, DMSO, etc.) | **E** | No additives |
| Manufacturer of plates/tubes and catalog number | D | ThermoFisher Scientific |
| Complete thermocycling parameters | **E** | 45°C for 20 min and 95°C for 10 min, followed by 45 cycles of 95°C for 15 s and 60°C for 1 min |
| Reaction setup (manual/robotic) | D | Manual set-up in a disinfected dead air box (10% bleach with fifteen minutes of contact time, UV for fifteen minutes, and a final cleaning step with 70% ethanol) |
| Manufacturer of qPCR instrument | **E** | ThermoFisher Scientfic |
| **qPCR VALIDATION** |  |  |
| Evidence of optimisation (from gradients) | D | See Liu *et al*. 2013^13^ and Liu *et al*. 2016^2^ |
| Specificity (gel, sequence, melt, or digest) | **E** | See Liu *et al*. 2013^13^ and Liu *et al*. 2016^2^ |
| Standard curves with slope and y-intercept | **E** | Table S2 |
| PCR efficiency calculated from slope | **E** | Table S2 |
| r2 of standard curve | **E** | Table S2 |
| Evidence for limit of detection | **E** | Table S2 |
| **DATA ANALYSIS** |  |  |
| qPCR analysis program (source, version) | **E** | QuantStudio Real-Time PCR Software V1.2 CDC |
| Cq method determination | **E** | Manual thresholding |
| Results of NTCs | **E** | We observed no amplification in our two PCR negative controls and two negative extraction control, except for the 16S assay (see results section). |
| Justification of number and choice of reference genes | **E** | N/A |
| Description of normalization method | **E** | Normalized to mass of stool |
| Software (source, version) | E | R Studio V2.2.2 |

**Text S6.** Positive and negative control results.

We extracted the 60 stool samples on two separate days and analyzed the extracts using TAC on two separate days. On each day of extractions, we included one negative extraction control and on each day of TAC analysis we included an additional PCR negative control. We observed no amplification in the pathogen targets among these four negative controls before a Cq of 35. Across the two days of TAC analysis, we included five PCR positive controls. The PCR positive controls amplified as expected, except for adenovirus 40/41 and hepatitis A virus, which were excluded from the analysis.

**Table S5.** Quantitative model parameters.

| **Model Variable** | **Parameters Used** | **References** |
| --- | --- | --- |
| **Infectious Unit Conversion** | | |
| Gene copy numbers per organism^1^ | *Acanthamoeba*: 600 copies per amoeba  *Cryptosporidium:* 20 copies per cyst  *Giardia:* 16 gene copies per cyst  EPEC (atypical): 1 gene copy per bacterium  EPEC (typical) / EAEC / ETEC: 1-16 gene copies per bacterium  *Helicobacter pylori*  *Blastocystis*  *Shigella*  *Plesiomonas shigelloides*  *Salmonella*  *Yersinia enterocolitica*  *Trichuris* | Yang *et al*. 1994^14^  McLauchlin *et al*. 1999^15^  Franzén *et al*. 2013^16^  Barbau-Piednoir et al. 2018^17^  Donnenberg *et al*. 1992^18^, Corsi *et al*. 2016^19^  Shukla *et al*. 2011^20^  Poirier *et al*. 2014^21^  Ashida and Sasakawa 2015^22^  Liu *et al*. 2016^2^  Liu *et al*. 2016^2^  Whatmore and Dowson 1999^23^  Pecson et al. 2006^24^ |
| Pathogen Concentration Per Gram Stool (for detects) |  | We divided gene copy concentrations by the number of gene copies per genome to estimate the number of pathogens per gram of stool |
| **Monte Carlo Simulation** | | |
| Non-detect values | 0 |  |
| *Acanthamoeba* number per gram stool in detects | mean = LN (2.67, 0.11)  sd = LN (0.29, 0.08) | MLE, this study |
| *Cryptosporidium* cyst number per gram stool in detects | mean = LN (5.62, 0.44)  sd = LN (0.99, 0.31) | MLE, this study |
| *Giardia* cyst number per gram stool in detects | mean = LN (5.02, 0.42)  sd = LN (0.84, 0.30) | MLE, this study |
| Atypical EPEC (eae) bacterium number per gram stool in detects | mean = LN (6.99, 0.30)  sd = LN (1.41, 0.21) | MLE, this study |
| Typical EPEC (bfpA) bacterium number per gram stool in detects | mean = LN (6.01, 0.46)  sd = LN (1.23, 0.33) | MLE, this study |
| Binomial distribution of *Acanthamoeba* detects | P(7/59) | This study |
| Binomial distribution of *Cryptosporidium* detects | P(5/59) | This study |
| Binomial distribution of *Giardia* detects | P(4/59) | This study |
| Binomial distribution of Typical Atypical EPEC (eae) detects | P(22/59) | This study |
| Binomial distribution of Atypical Typical EPEC (bfpA) detects | P(7/59) | This study |
| Weekly Reduction in Stools by Pit Stop intervention (Inner loop) | N = 18 | Amato et al. 2022^25^ |
| Number of grams per stool | 106 g | Cummings *et al*. 1992^26^, Berendes *et al*. 2018^27^ |
| Weeks per Year (2^nd^ loop) | N = 52 |  |
| Total Annual Simulations (Outer Loop) | N = 1000 |  |

**Table S6.** Quantal detection data, all targets.

| **No.** | **Pathogen** | **All discarded feces (n=59)** | **Human feces (n=12)** | **Non-human feces (n=47)** |
| --- | --- | --- | --- | --- |
| **Bacteria** |  |  |  |  |
| 1 | *EPEC^1^ (atypical)* | 37% | 42% | 36% |
| 2 | *EPEC^1^ (typical)* | 12% | 0% | 15% |
| 3 | *EAEC^2^ (AAIC/AATA)* | 3% | 8% | 2% |
| 4 | *ETEC^3^ (LT/ST)* | 3% | 8% | 2% |
| 5 | *H. pylori* | 3% | 17% | 0% |
| 6 | *Shigella spp. /EIEC (ipaH)* | 2% | 8% | 0% |
| 7 | *Plesiomonas shigelloides* | 2% | 8% | 0% |
| 8 | *Salmonella spp.* | 2% | 0% | 2% |
| 9 | *Yersinia enterocolitica* | 2% | 8% | 0% |
| 10 | *Campylobacter jejuni/coli* | 0% | 0% | 0% |
| 11 | *Clostridium diff.* | 0% | 0% | 0% |
| 12 | *E. coli_O157_H7* | 0% | 0% | 0% |
| 13 | *STX (I/II)* | 0% | 0% | 0% |
| **Viruses** |  |  |  |  |
| 1 | *Norovirus (GI/II)* | 2% | 0% | 2% |
| 2 | *Adenovirus 40/41* | 0% | 0% | 0% |
| 3 | *Astrovirus* | 0% | 0% | 0% |
| 4 | *Hepatitis A Virus* | 0% | 0% | 0% |
| 5 | *Rotavirus* | 0% | 0% | 0% |
| 6 | *Sapovirus (I/II/IV/V)* | 0% | 0% | 0% |
| 7 | *SARS-CoV-2* | 0% | 0% | 0% |
| **Protozoa** |  |  |  |  |
| 1 | *Acanthamoeba spp.* | 12% | 17% | 11% |
| 2 | *Cryptosporidium spp.* | 8% | 0% | 11% |
| 3 | *Giardia spp.* | 7% | 0% | 9% |
| 4 | *Blastocystis spp.* | 3% | 8% | 2% |
| 5 | *Balantidium coli* | 2% | 0% | 2% |
| 6 | *Cystoisospora belli* | 0% | 0% | 0% |
| 7 | *Cyclospora cayetanensis* | 0% | 0% | 0% |
| 8 | *E. bieneusi* | 0% | 0% | 0% |
| 9 | *E. intestinalis* | 0% | 0% | 0% |
| 10 | *Entamoeba hystolytica* | 0% | 0% | 0% |
| 11 | *Entamoeba spp.* | 0% | 0% | 0% |
| **STHs** |  |  |  |  |
| 1 | *Trichuris spp.* | 3% | *8% | 2% |
| 2 | *Ancyclostoma* | 0% | 0% | 0% |
| 3 | *Ascaris spp.* | 0% | 0% | 0% |
| 4 | *E. vermicularis* | 0% | 0% | 0% |
| 5 | *Hymenolepis nana* | 0% | 0% | 0% |
| 6 | *Necator americanus* | 0% | 0% | 0% |
| 7 | *Strongyloides* | 0% | 0% | 0% |
| **mtDNA** |  |  |  |  |
|  | hCYTB484 (human specific marker) | 22% | 100% |  |
| ^1^Enteropathogenic *Escherichia coli*  ^2^Enteroaggregative *Escherichia coli*  ^3^Enterotoxigenic *Escherichia coli*  *No parasites seen in human mtDNA-positive^5^ (Text S5) *Trichuris* sample on microscopy or subsequent qPCR analysis | | | | |

**Figure S1**. Intervention and stool sample locations, central San Francisco, CA, USA. Background map source: maps.google.com.

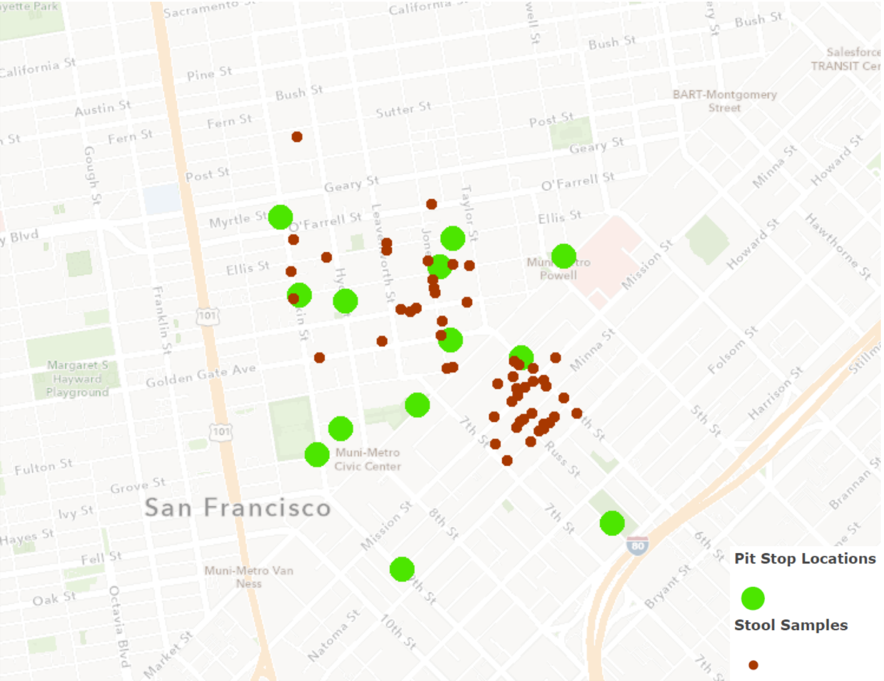

**Figure S2**. Intervention toilet in the Tenderloin, San Francisco. Photo credit: Jay Graham

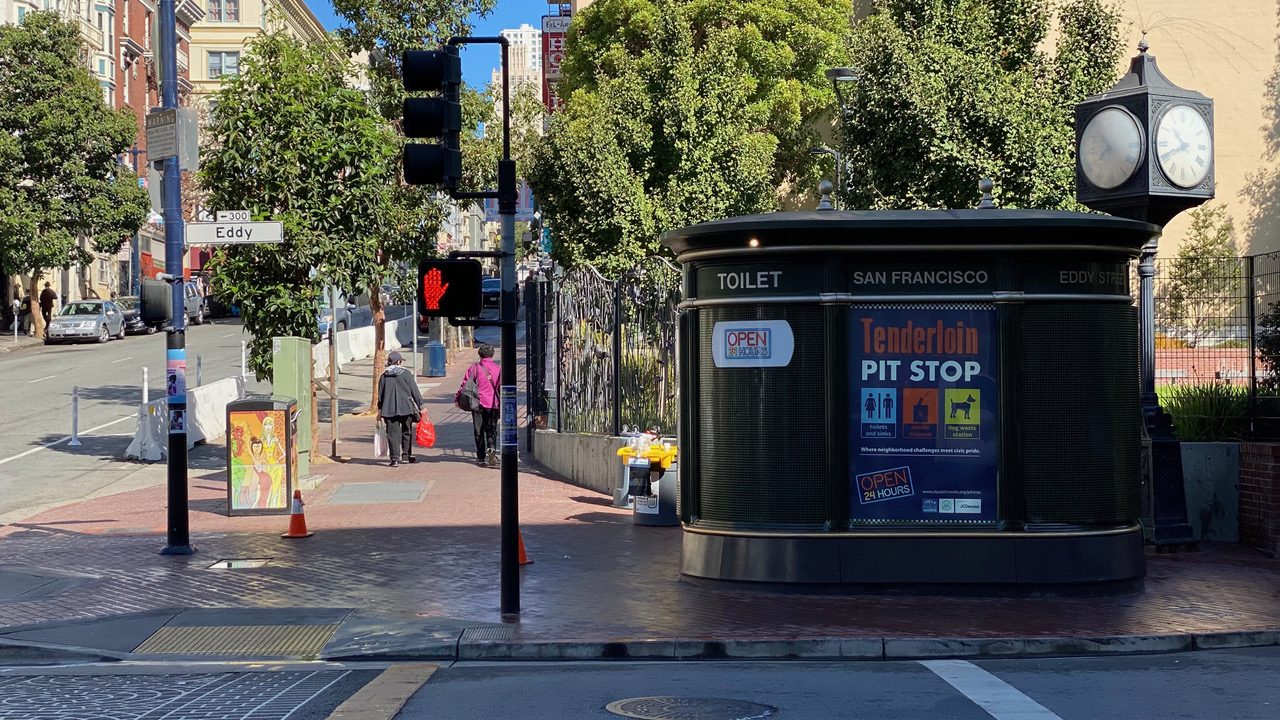

**Figure S3.** Scatterplot (top) and box plot (bottom) showing log_10_ gene copies of mtDNA using hCYTB484 assay^5^, normalized by dsDNA concentration. Human and non-human stools had clear visual groupings in terms of normalized concentration, with human samples grouped near the top of the plot.

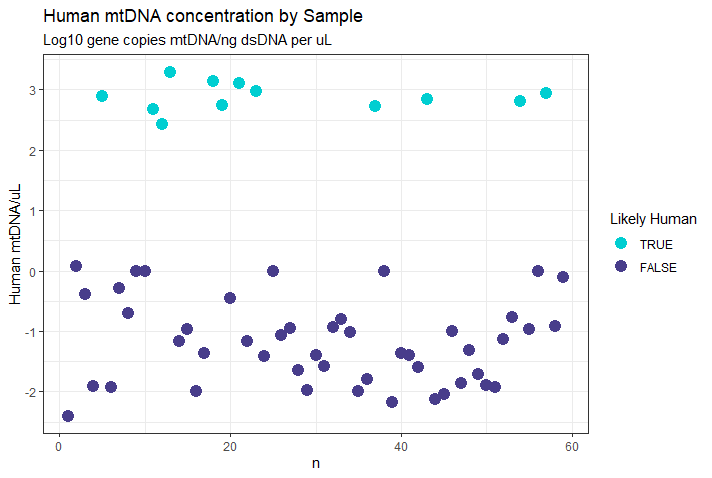

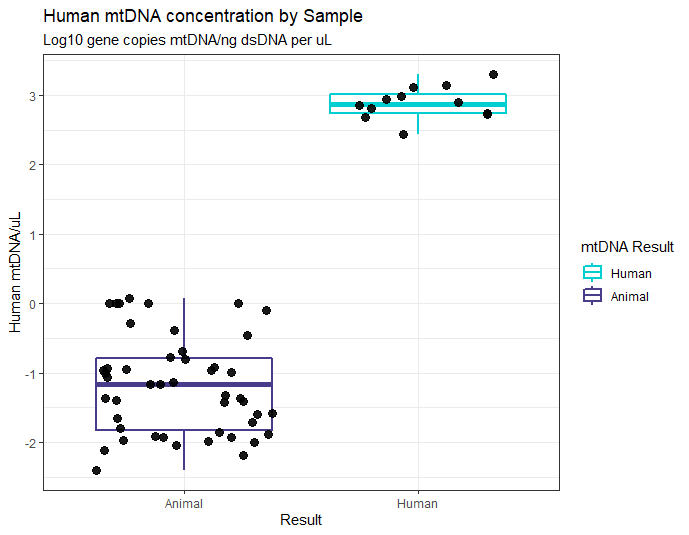

**Figure S4.** Microscopy images from mtDNA negative *Trichuris* sample. From top to bottom: *Trichuris vulpis, Toxocara canis,* hookworm.

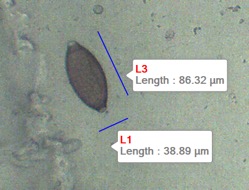

*Trichuris vulpis* (67 ova per gram found)

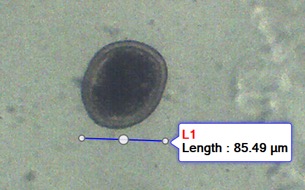

*Toxocara canis* (950 ova per gram found)

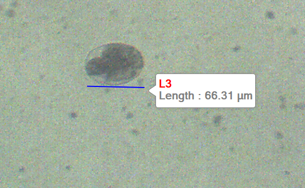

Hookworm, species unknown, (5481 ova per gram found)

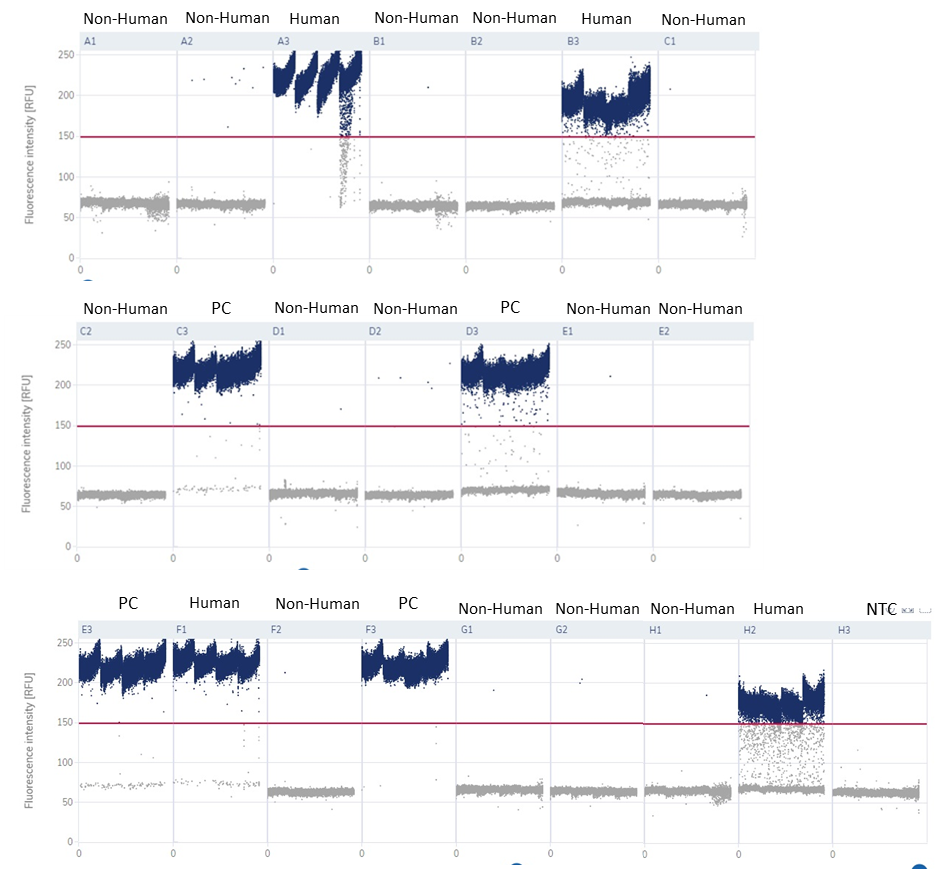
**Figure S5. mtDNA dPCR RFU plots** demonstrating partition classification (i.e., threshold separating positive and negative bands) in a plate with human and non-human samples and controls

Note: PC = positive control

**Figure S6. Amplification and multicomponent plots**

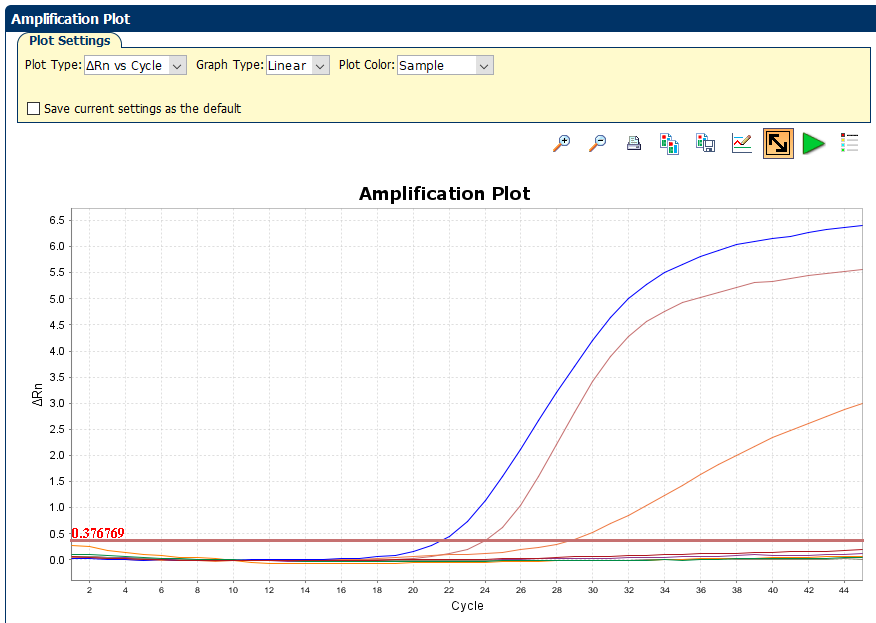

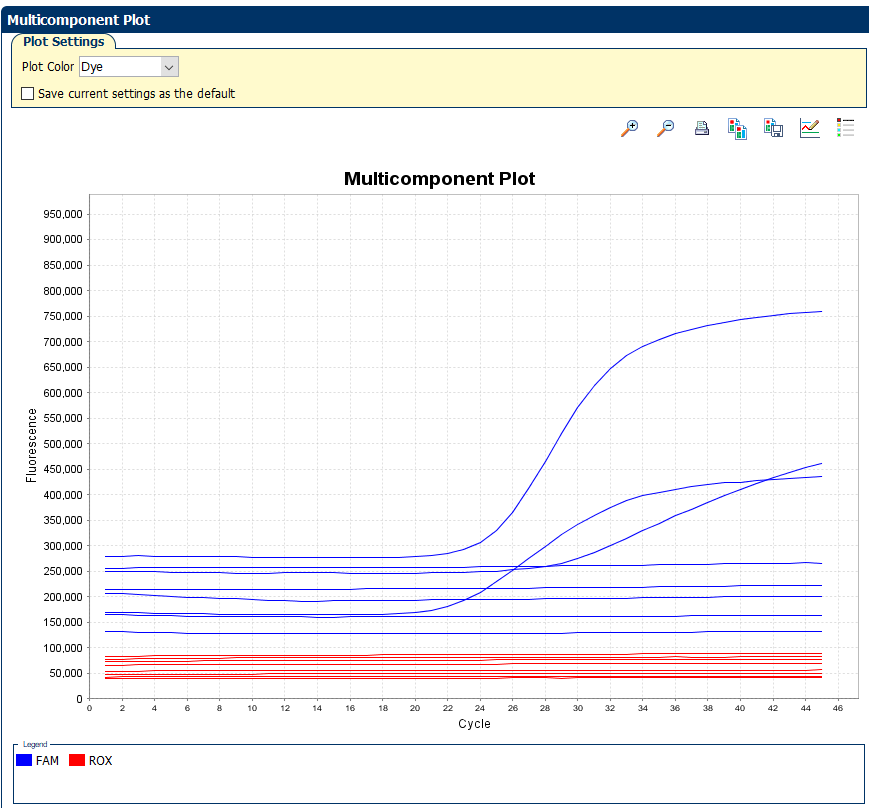

**Table S7.** dMIQE checklist for mtDNA dPCR

| ITEM TO CHECK | PROVIDED | COMMENT |
| --- | --- | --- |
|  | Y/N |  |
| 1. SPECIMEN |  |  |
| Detailed description of specimen type and numbers | Y | Found in Sample collection (type) and results (numbers) in manuscript |
| Sampling procedure (including time to storage) | Y | Found in the Sample collection section in the manuscript |
| Sample aliquotation, storage conditions and duration | Y | Found in Sample collection and sample preparation section of manuscript |
| 2. NUCLEIC ACID EXTRACTION |  |  |
| Description of extraction method including amount of sample processed | Y | Found in Sample preparation section in the manuscript |
| Volume of solvent used to elute/resuspend extract | Y | Text S2 (100µL) |
| Number of extraction replicates | N/A | None tested using dPCR |
| Extraction blanks included? | N/A | None tested using dPCR |
| 3. NUCLEIC ACID ASSESSMENT AND STORAGE |  |  |
| Method to evaluate quality of nucleic acids | N/A | Not done |
| Method to evaluate quantity of nucleic acids (including molecular weight and calculations when using mass) | Y | Sample analysis: qubit (see “sample preparation and analysis” in manuscript) |
| Storage conditions: temperature, concentration, duration, buffer, aliquots | Y | Text S1-S3 of the supplementary information |
| Clear description of dilution steps used to prepare working DNA solution | Y | Text S5, template was undiluted |
| 4. NUCLEIC ACID MODIFICATION | NA | N/A, not performed |
| 5. REVERSE TRANSCRIPTION | NA | All templates measured with dPCR were DNA templates |
| 6. dPCR OLIGONUCLEOTIDES DESIGN AND TARGET INFORMATION |  |  |
| Sequence accession number or official gene symbol | Y | hCYTB484 |
| Method (software) used for design and in silico verification | Y | Zhu, K.; Suttner, B.; Pickering, A.; Konstantinidis, K. T.; Brown, J. A Novel Droplet Digital PCR Human MtDNA Assay for Fecal Source Tracking. Water Res. 2020, 183, 116085.https://doi.org/10.1016/J.WATRES.2020.116085.^5^ |
| Location of amplicon | Y | Zhu, K.; Suttner, B.; Pickering, A.; Konstantinidis, K. T.; Brown, J. A Novel Droplet Digital PCR Human MtDNA Assay for Fecal Source Tracking. Water Res. 2020, 183, 116085. https://doi.org/10.1016/J.WATRES.2020.116085.^5^ |
| Amplicon length | Y | Text S5 |
| Primer and probe sequences (or amplicon context sequence)** | Y | Text S5 |
| Location and identity of any modifications | N/A | None in this study |
| Manufacturer of oligonucleotides | Y | Text S5 |
| 7. dPCR PROTOCOL |  |  |
| Manufacturer of dPCR instrument and instrument model | Y | Text S5 |
| Buffer/kit manufacturer with catalogue and lot number | Y | QIAcuity Probe PCR Kit (5 ml)  Cat. No. / ID: 250102 |
| Primer and probe concentration | Y | Text S5 |
| Pre-reaction volume and composition (incl. amount of template and if restriction enzyme added) | Y | Text S5 |
| Template treatment (initial heating or chemical denaturation) | N/A | No template treatment |
| Polymerase identity and concentration, Mg++ and dNTP concentrations*** | N/A | Proprietary, (Qiagen, Hilden, Germany) |
| Complete thermocycling parameters | Y | Text S5 |
| 8. ASSAY VALIDATION |  |  |
| Details of optimization performed | N/A | Performed by Zhu *et al*. 2020^5^ |
| Analytical specificity (vs. related sequences) and limit of blank (LOB) | N/A | Performed by Zhu *et al*. 2020^5^ |
| Analytical sensitivity/LoD and how this was evaluated | N/A | Performed by Zhu *et al*. 2020^5^ |
| Testing for inhibitors (from biological matrix/extraction) | N/A | Did not do this |
| 9. DATA ANALYSIS |  |  |
| Description of dPCR experimental design | Y | Sample Preparation and analysis, Text S5 |
| Comprehensive details negative and positive of controls (whether applied for QC or for estimation of error) | Y | Text S5 |
| Partition classification method (thresholding) | Y | Text S5 |
| Examples of positive and negative experimental results (including fluorescence plots in supplemental material) | Y | Figure S5 |
| Description of technical replication | Y | Text S5, Figure S5 |
| Repeatability (intra-experiment variation) | Y | Text S5 |
| Reproducibility (inter-experiment/user/lab etc. variation ) | N |  |
| Number of partitions measured (average and standard deviation ) | Y | Text S5: Average partitions measured were 24879 (standard deviation = 1590). |
| Partition volume | N | Calculated by software |
| Copies per partition (λ or equivalent ) (average and standard deviation) | Y | Text S5 |
| dPCR analysis program (source, version) | Y | Text S5 |
| Description of normalisation method | Y | Sample preparation and analysis: “We normalized mtDNA gene copy estimates to ng of dsDNA, and compared results against values reported in the literature.” |
| Statistical methods used for analysis | Y | Sample preparation and analysis: “We normalized mtDNA gene copy estimates to ng of dsDNA, and compared results against values reported in the literature.” |
| Data transparency | Raw data available on request | Raw data available on request |

**References: Supporting Information**

1.	Kodani, M.; Winchell, J. M., Engineered combined-positive-control template for real-time reverse transcription-PCR in multiple-pathogen-detection assays. *J Clin Microbiol* **2012,** *50*, (3), 1057-60.

2.	Liu, J.; Gratz, J.; Amour, C.; Nshama, R.; Walongo, T.; Maro, A.; Mduma, E.; Platts-Mills, J.; Boisen, N.; Nataro, J.; Haverstick, D. M.; Kabir, F.; Lertsethtakarn, P.; Silapong, S.; Jeamwattanalert, P.; Bodhidatta, L.; Mason, C.; Begum, S.; Haque, R.; Praharaj, I.; Kang, G.; Houpt, E. R., Optimization of Quantitative PCR Methods for Enteropathogen Detection. *PLOS ONE* **2016,** *11*, (6), e0158199.

3.	Cringoli, G.; Maurelli, M. P.; Levecke, B.; Bosco, A.; Vercruysse, J.; Utzinger, J.; Rinaldi, L., The Mini-FLOTAC technique for the diagnosis of helminth and protozoan infections in humans and animals. *Nature Protocols* **2017,** *12*, (9), 1723-1732.

4.	Barda, B. D.; Rinaldi, L.; Ianniello, D.; Zepherine, H.; Salvo, F.; Sadutshang, T.; Cringoli, G.; Clementi, M.; Albonico, M., Mini-FLOTAC, an innovative direct diagnostic technique for intestinal parasitic infections: experience from the field. *PLoS Negl Trop Dis* **2013,** *7*, (8), e2344.

5.	Zhu, K.; Suttner, B.; Pickering, A.; Konstantinidis, K. T.; Brown, J., A novel droplet digital PCR human mtDNA assay for fecal source tracking. *Water Res* **2020,** *183*, 116085.

6.	Rudko, S. P.; Ruecker, N. J.; Ashbolt, N. J.; Neumann, N. F.; Hanington, P. C., Enterobius vermicularis as a Novel Surrogate for the Presence of Helminth Ova in Tertiary Wastewater Treatment Plants. *Appl Environ Microbiol* **2017,** *83*, (11).

7.	Kann, S.; Hartmann, M.; Alker, J.; Hansen, J.; Dib, J. C.; Aristizabal, A.; Concha, G.; Schotte, U.; Kreienbrock, L.; Frickmann, H., Seasonal Patterns of Enteric Pathogens in Colombian Indigenous People-A More Pronounced Effect on Bacteria Than on Parasites. *Pathogens* **2022,** *11*, (2).

16.	Franzén, O.; Jerlström-Hultqvist, J.; Einarsson, E.; Ankarklev, J.; Ferella, M.; Andersson, B.; Svärd, S. G., Transcriptome profiling of Giardia intestinalis using strand-specific RNA-seq. *PLoS Comput Biol* **2013,** *9*, (3), e1003000.

22.	Ashida, H.; Sasakawa, C., Shigella IpaH Family Effectors as a Versatile Model for Studying Pathogenic Bacteria. *Front Cell Infect Microbiol* **2015,** *5*, 100.

23.	Whatmore, A. M.; Dowson, C. G., The autolysin-encoding gene (lytA) of Streptococcus pneumoniae displays restricted allelic variation despite localized recombination events with genes of pneumococcal bacteriophage encoding cell wall lytic enzymes. *Infect Immun* **1999,** *67*, (9), 4551-6.

24.	Pecson, B. M.; Barrios, J. A.; Johnson, D. R.; Nelson, K. L., A real-time PCR method for quantifying viable ascaris eggs using the first internally transcribed spacer region of ribosomal DNA. *Appl Environ Microbiol* **2006,** *72*, (12), 7864-72.

25.	Amato, H. K.; Martin, D.; Hoover, C. M.; Graham, J. P., Somewhere to go: assessing the impact of public restroom interventions on reports of open defecation in San Francisco, California from 2014 to 2020. *BMC Public Health* **2022,** *22*, (1), 1673.

26.	Cummings, J. H.; Bingham, S. A.; Heaton, K. W.; Eastwood, M. A., Fecal weight, colon cancer risk, and dietary intake of nonstarch polysaccharides (dietary fiber). *Gastroenterology* **1992,** *103*, (6), 1783-9.

27.	Berendes, D. M.; Yang, P. J.; Lai, A.; Hu, D. L.; Brown, J., Estimation of global recoverable human and animal faecal biomass. *Nature Sustainability* **2018,** *1*, 679-685.
